# Sufficient reproduction numbers to prevent recurrent epidemics

**DOI:** 10.1101/2024.02.15.24302902

**Authors:** Lorenzo Mari, Cristiano Trevisin, Andrea Rinaldo, Marino Gatto

## Abstract

Current practice in the design and evaluation of epidemic control measures, including vaccination, is largely based on reproduction numbers (RNs), which represent prognostic indexes of long-term disease transmission, both in naïve populations (basic RN) and in the presence of prior exposure or interventions (effective RN). A standard control objective is to establish herd immunity, e.g., by immunizing enough susceptible individuals to achieve RN<1. However, attaining this goal is not sufficient to avoid transient outbreaks that, in the short term, might revamp epidemics by coalescence of subthreshold flare-ups.
Using reactivity analysis applied to a discrete SIR model with age-of-infection structure, we determine sufficient conditions to prevent transient epidemic dynamics and recurrent, non-periodic outbreaks due to imported cases. These conditions are based on fundamental infection characteristics, namely the average infectiousness clearance rate, the generation time distribution, and the RN.
We show that preventing subthreshold epidemicity requires stricter RN thresholds than simply maintaining RN< 1. Taking into account a wide spectrum of respiratory viral infections, epidemicity-curbing RN thresholds vary between 0.10 (rubella) and 0.51 (MERS), with a median of 0.26 close to the estimate of 0.24 for the ancestral SARS-CoV-2 virus. The portion of the population that needs to be included in containment efforts to avoid short-term outbreaks is considerably higher than herd immunity thresholds (HITs) based solely on the basic RN (e.g., 93% vs. 72% for ancestral SARS-CoV-2). We also find that subthreshold epidemicity is harder to prevent for pathogens with a longer mean generation time, smaller standard deviation of the generation time distribution, longer duration of infection, and higher RN.
Determining sufficient RN thresholds to prevent transient outbreaks is a key challenge in disease ecology, with practical consequences for the design of control measures, as the weaker RN reductions and HITs associated with customary control targets may prove ineffective in preventing potentially recurrent flare-ups. Due to its modest data requirements, our modeling framework may also have important implications for human and non-human diseases caused by emerging pathogens.

**Data/Code:** This work has no associated primary data. The secondary data used to parameterize the model are described in Supporting Information (SI, section S1; see also SI References). The main code to perform discrete epidemicity analysis has been uploaded for review purposes; it will be made available on a public repository once the editorial process is completed.

## Introduction

During the COVID-19 pandemic, reproduction numbers (RNs), in their basic (*ℛ*_0_; Heesterbeek, 2002), control (*ℛ*_c_; Brauer, 2008), or effective (*ℛ*_*t*_; Nishiura and Chowell, 2009) formulations, have become the currency of communication as their updated values were continuously reported as part of the day-by-day messages of hope or despair (Cinelli et al., 2020). After seeding of the infection within a community, herd immunity is reached if/when the RN remains permanently below the unit threshold, by means of suitable containment measures (typically sufficient vaccinations) and/or acquired disease-specific immunity (Brett and Rohani, 2020). However, herd immunity, possibly affected by population heterogeneity (Britton et al., 2020), is not sufficient to prevent subthreshold (Van den Driessche and Watmough, 2002; Mari et al., 2019) bursts of epidemic activity (i.e., occurring with RN < 1), nor guarantee against stuttering transmission chains (Lloyd-Smith et al., 2009; Blumberg and Lloyd-Smith, 2013) or wavelets (Callaway, 2023). Under appropriate conditions, a certain number of cases imported into a population in which vaccinations or other containment measures maintain below unity the effective RN may still spur transient increases in infection prevalence. Although possibly small individually, recurrent subthreshold outbreaks can merge into larger ones (Figure 1). Therefore, preventing pathogens from even temporarily spreading within a population gains importance because case imports are often unavoidable, as testified, for example, by MERS-CoV outbreaks in the Middle East (Gardner and MacIntyre, 2014) or measles in the US that occur despite control efforts (Blumberg et al., 2015). The issue of recurrent epidemics has been explored via deterministic and stochastic models, which have highlighted their various features, often concentrating on seasonal dynamics (Johansen, 1996; Finkenstädt et al., 2002; Stone et al., 2007; Begon et al., 2009; Zheng et al., 2015; Cao et al., 2019; Saad-Roy et al., 2020).

**Figure 1:**
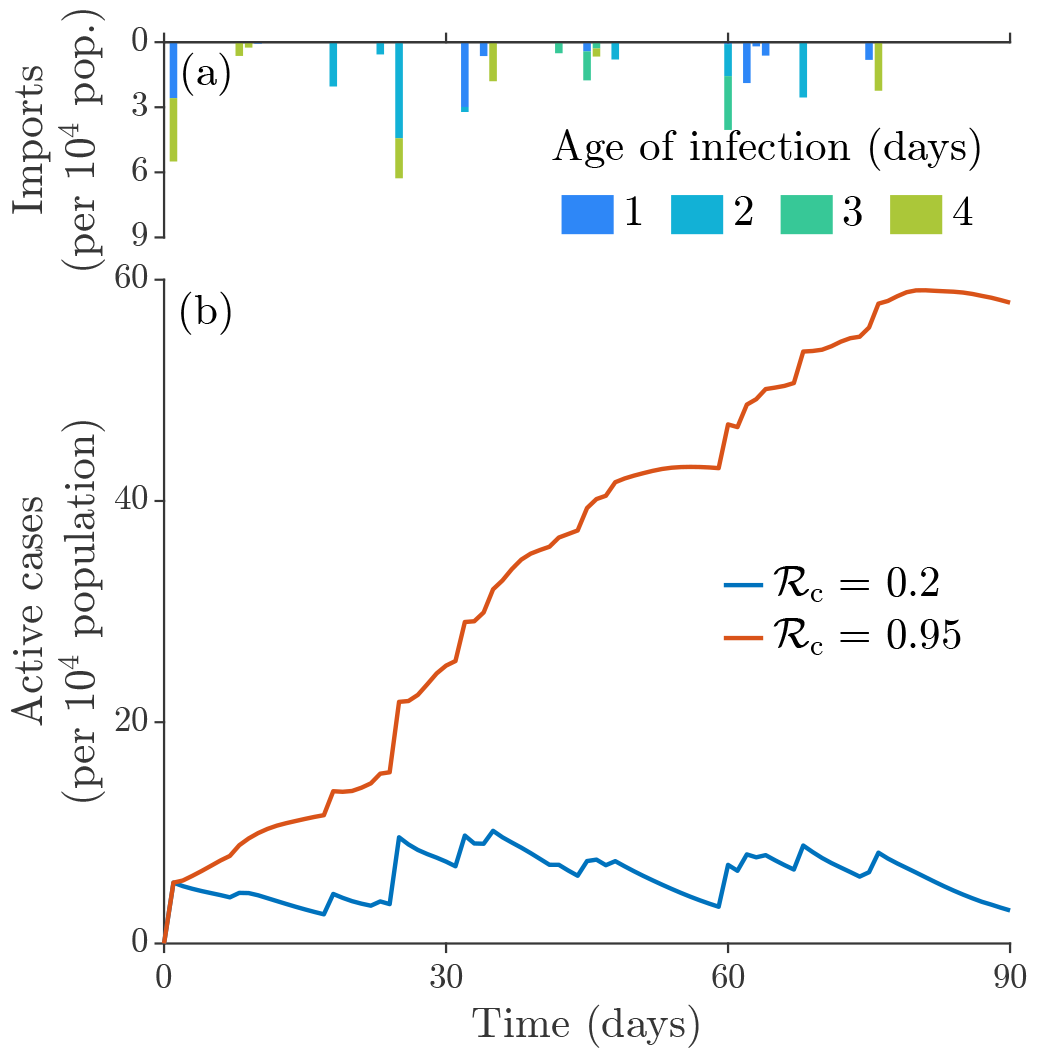
Examples of subthreshold epidemiological dynamics within a population in the presence of recurrent imports of infections. (a) Time series of imported cases, generated by assuming that small clusters of infections (one case per 10^4^ population on average) are imported recurrently (two times per week on average) into the population. Both the intensity and the timing of the imports are drawn from exponential probability distributions (note that multiple events can occur on the same day). The age of infection of the imported cases is randomly assigned for each cluster using a discrete uniform distribution with support spanning from one to four days, mimicking a latent infection phase during which infected people are more likely to travel. (b) Prevalence of infection within an initially naïve population simulated with the discrete-time transmission model with age-of-infection structure (Materials and Methods). The depletion of the susceptible pool is assumed to be negligible, therefore the effective RN is approximated with a constant control RN, *ℛ*_c_. Recurrent case imports do not generate transient epidemic outbreaks with strong containment (*ℛ*_c_ = 0.2, blue curve), while they trigger subthreshold transmission chains eventually coalescing into a larger outbreak with weaker interventions (*ℛ*_c_ = 0.95, red curve), which would anyway prevent endemicity in the long run. Projections are made under the assumption of gamma-distributed generation times (with an average of 5.2 days and a standard deviation of 1.7 days) and a total duration of the infection of 11.7 days, inclusive of the latent phase (corresponding to the parameterization for the ancestral strain of SARS-CoV-2; see Table 1).

Our goal here is to establish a methodological framework that allows to formally determine whether by suitably reducing the RN well below unity it is possible to obtain not only herd immunity but also epidemicity immunity, that is, fostering conditions conducive to the avoidance of transient epidemics. It should be noted that for many infectious diseases for which vaccines are available, the fraction of people who need to be vaccinated is often empirically set at a level larger than the herd immunity threshold HIT = 1 − 1*/*ℛ_0_. In other words, a sort of practical precautionary principle is applied with the aim of achieving more than simple herd immunity for diseases that are deemed particularly dangerous to human health. Our framework can help establish an exact quantification of that principle by exploring how much effort is needed to achieve the goal of avoiding transient, possibly recurrent epidemics.

**Table 1:** Basic parameters for the evaluation of discrete epidemicity applied to 15 examples of infectious diseases caused by respiratory viruses. AV and SD are the average value and the standard deviation of the generation times, assumed to be gamma-distributed, while *γ* is the instantaneous exit rate from the infected compartment (whose inverse corresponds, as a first approximation, to the average duration of infection; note that the estimates for COVID-19, MERS, SARS, and smallpox also account for disease-related mortality). See SI (section S1) for details on each parameterization and the relevant literature references.

| Disease | AV (days) | SD (days) | $1/\gamma$ (days) | $\mathcal{R}_0$ |
| --- | --- | --- | --- | --- |
| Adenovirus infection | 7.1 | 2.6 | 11.5 | 5.1 |
| COVID-19 (ancestral strain) | 5.2 | 1.7 | 11.7 | 3.6 |
| Influenza A(H1N1)pdm09 | 2.8 | 1.0 | 3.6 | 1.18 |
| Influenza A(H3N2) | 2.3 | 0.80 | 4.5 | 1.16 |
| Influenza B | 3.7 | 2.0 | 6.2 | 1.24 |
| MERS | 6.8 | 4.1 | 10.8 | 0.7 |
| Measles | 11.7 | 2.5 | 14 | 14.5 |
| Mumps | 18.0 | 3.5 | 31 | 4.28 |
| Parainfluenza | 5.2 | 2.2 | 9.8 | 2.7 |
| Pertussis | 22.8 | 6.4 | 28 | 5.5 |
| RSV | 7.5 | 2.1 | 13 | 1.32 |
| Rubella | 18.3 | 2.0 | 20.1 | 5.9 |
| SARS | 10.0 | 2.8 | 10 | 3.0 |
| Smallpox | 17.6 | 3.2 | 23.2 | 4.5 |
| Varicella | 14.0 | 2.4 | 19 | 6.73 |

While the theory behind the long-term growth of epidemic trajectories is well established, the formal study of transient short-term dynamics in epidemiology and disease ecology has received much less attention. Recently, the concept of epidemicity (Hosack et al., 2008; Mari et al., 2018, 2019, 2021; Trevisin et al., 2022), based on ecological reactivity (Neubert and Caswell, 1997) and its generalizations (Mari et al., 2017), has been proposed for continuous-time epidemiological systems as a tool to analyze the short-term dynamics of epidemic trajectories that are bound to fade in the long run because RN < 1. In that context, epidemicity formally quantifies the initial instantaneous growth rate of the fastest-growing perturbation to an asymptotically stable disease-free equilibrium (DFE). Clearly, avoiding recurrent outbreaks triggered, e.g., by the repeated import of infections requires guaranteeing that no perturbations can be amplified over time. Therefore, we can define an epidemic as non-recurrent if the prevalence of infection always decreases after the import of a cluster of infected cases, independently of their age of infection or other epidemiological traits.

To more closely resemble the discrete-time nature of surveillance data that is typically used to estimate RNs (Cori et al., 2013; Liu et al., 2018; Gostic et al., 2020; Zhang et al., 2020; Cereda et al., 2021; Trevisin et al., 2023), we adopt a susceptible-infected-removed model with age-of-infection structure (Kermack and McKendrick, 1927; Cao et al., 2019) and discretize it in time and age of infection (Allen and van den Driessche, 2008), thereby obtaining an epidemiological projection matrix that can be used to simulate the transmission process. The projection matrix can be made time- or policy-dependent by substituting the basic RN with the effective or control RNs (Materials and Methods). We assume that RNs can be estimated in real-time from data (Cori et al., 2013; Liu et al., 2018; Gostic et al., 2020; Zhang et al., 2020; Cereda et al., 2021; Trevisin et al., 2023), incorporated into the projection matrix, and used to forecast whether the intensity of the epidemic can grow over time. The application of the model requires some basic knowledge about the main epidemiological characteristics of the pathogen under study, namely the average removal time from the infected compartment (due to recovery or death), the distribution of disease generation times, and the basic RN, as shown in Table 1 for a selection of 15 respiratory viral infections (Leung, 2021). The details on the parameterization of each infectious disease are reported in the Supporting Information (SI, section S1).

## Materials and Methods

### Continuos-time/age infection process

We describe the spread of a viral respiratory virus (Leung, 2021) within a well-mixed population using a susceptible-infected-removed (SIR) transmission process based on a Kermack-McKendrick-type set of integrodifferential equations (Kermack and McKendrick, 1927; Nishiura and Chowell, 2009), namely

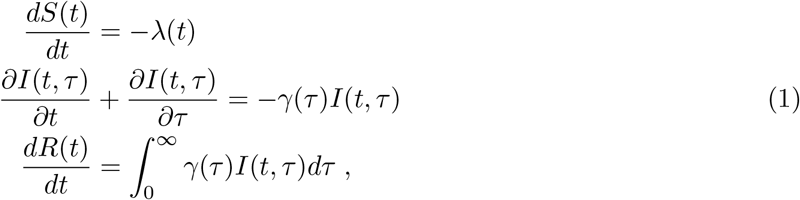

where:

- *S*(*t*), *I*(*t, τ*), and *R*(*t*) are the abundances of susceptible, infected, and removed individuals at time *t* (and age of infection *τ* for infected individuals);
- *γ*(*τ*) is the instantaneous removal rate from the infected compartment for individuals with age of infection *τ* because of recovery or death;
- 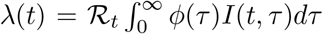 is the incidence of new cases, with *ℛ*_*t*_ and *ϕ*(*τ*) being respectively the effective reproduction number (RN), defined as the number of secondary cases produced by one index case throughout the duration of the infection, and the secondary infectivity rate (per unit of RN) of individuals with age of infection *τ*. The latter quantity is related to the probability distribution *β*(*τ*) of generation times, defined as the time intervals between subsequent infection events in a transmission chain, via the relationship *β*(*τ*) = *ϕ*(*τ*)*p*(*τ*), where 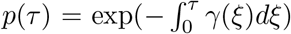 is the probability of still being in the infected compartment at age of infection *τ*.

Model (1) is subject to the initial conditions *S*(0) = *s*_0_, *I*(0, *τ*) = *i*_0_(*τ*), and *R*(0) = *r*_0_, as well as to the boundary condition *I*(*t*, 0) = *λ*(*t*). Given our focus on transient dynamics, we consider neither demographic (birth, death) nor epidemiological (loss of acquired immunity) processes which are expected to operate over longer timescales.

It can be noticed that model (1) is linear in the state variables. However, this is not an inherent limitation of the mathematical framework. In fact, the effective RN can be used to summarize the effects of both intrinsic (decline in susceptible individuals) and extrinsic (implementation of control measures) factors of time-dependent changes in disease transmission (Nishiura and Chowell, 2009). If the depletion of the susceptible pool can be assumed to be negligible (e.g., at the beginning of an outbreak) and the containment effort does not change over the temporal interval of interest, the effective RN can be replaced by the control RN (*ℛ*_c_; Brauer, 2008) or, in the absence of controls, by the basic RN (*ℛ*_0_; Heesterbeek, 2002).

### Time- and age-discrete version of the infection process

Epidemiological surveillance can produce information only as a discrete-time stream (e.g., daily or weekly bulletins). Model (1) can be discretized appropriately in both time and age of infection to better adapt to the available data. Without loss of generality, we apply a daily step for both independent variables, as often done in applications based on discrete-time modeling (Cori et al., 2013; Liu et al., 2018; Gostic et al., 2020; Zhang et al., 2020; Cereda et al., 2021; Trevisin et al., 2023). Also, we only focus on the infection subsystem identified by the infected compartment *I*(*t, τ*), which is basically decoupled from susceptible and removed individuals in model (1) if we assume, as we do, that the effective RN is known (in practice, it must be estimated independently from data). The discretized dynamics of the model reads

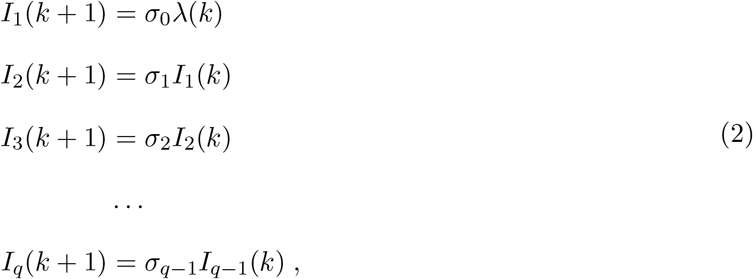

where:

- 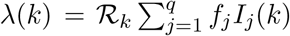 is the discrete version of the incidence function at day *k*, with 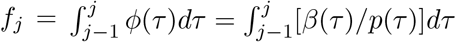 (in which the definite integral has typically to be approximated through a numerical integration formula, such as the trapezoid rule) and *q* being the maximum considered age of infection;
- *σ*_*j*−1_ = *p*(*j*)*/p*(*j* − 1), with *j* ∈ {1, …, *q*}, is the probability of remaining within the infected compartment for one more timestep (i.e., a day) for individuals with (discrete) age of infection *j*− 1.

Note that the fundamental property 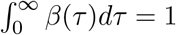 translates to 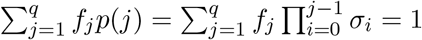, with *q* chosen so that *f*_*q*_ → 0 and *p*(*q*) → 0. Specifically, we select *q* so that the probability of being still infected after *q* days does not exceed a given threshold *x* and the integral of the generation time distribution up to *q* is not smaller than 1 − *x*, with *x* = 10^−4^.

In matrix notation, the discretized system (2) becomes **I**(*k* + 1) = **L**(*k*)**I**(*k*), with

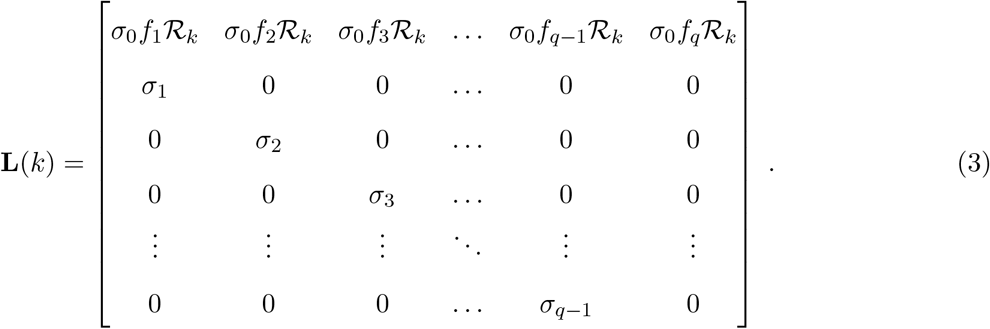

Matrix (3) has the structure of a so-called Leslie projection matrix (Leslie, 1945), perhaps the most fundamental tool in (age-structured) demography and population dynamics. The time dependency introduced by the presence of the effective RN allows the projection matrix to incorporate the temporal changes in susceptible prevalence and sanitary policy. As noted for the continuous model (1), if the depletion of the susceptible pool can be considered negligible and the control effort does not change over time, the effective RN can be replaced by the control/basic RN (depending on presence/absence of containment measures); in this case, the projection matrix and the associated system of difference equations become time-invariant.

### Long-term epidemiological dynamics

Using basic demographic theory (Caswell, 2000), it is immediate to determine that any epidemic trajectory will asymptotically continue to grow if 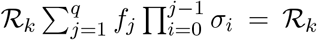 remains permanently above unity. Analogously, by analogy with the definition of *ℛ*_0_, one can prove that *ℛ*_*k*_ coincides with the spectral radius of the next-generation matrix **Q**(*k*) = **F**(*k*)[**Id** − **T**(*k*)]^−1^ (as defined for discrete-time systems; Allen and van den Driessche, 2008), where **F**(*k*) is a possibly time-variant matrix of new infections (a square matrix of size *q* with the first row corresponding to that of matrix (3), and all other elements being null), **Id** is the identity matrix, and **T**(*k*) = **L**(*k*) − **F**(*k*) is the transition matrix; therefore, *ℛ*_*k*_ > 1 is a sufficient (albeit not always necessary) condition for the epidemic to unfold in the long run (Dushoff et al., 1998; Van den Driessche and Watmough, 2002). In practice, the asymptotic nature of these results is often relaxed, as in the case of real-time approaches for the estimation of the effective RN (Cori et al., 2013; Liu et al., 2018; Gostic et al., 2020; Zhang et al., 2020; Cereda et al., 2021; Trevisin et al., 2023), in which a stream of surveillance data is assimilated to produce updated projections of the development of an outbreak over finite timescales of epidemiological interest, using a quasi-equilibrium approach. In this case, RN > 1 should be interpreted as a sufficient condition for an increase in reported cases during the current observation period.

### A theory of discrete epidemicity

To define a discrete epidemicity index, we resort to reactivity theory in its extension to discrete-time systems. For this family of problems, reactivity has been defined as (the natural logarithm of) the maximum growth achieved in one step by any perturbation to an asymptotically stable steady state (Caswell and Neubert, 2005): if at least some perturbations can be initially amplified, the equilibrium is defined as reactive. Epidemicity (that is, reactivity in the context of epidemiological applications) (Hosack et al., 2008; Mari et al., 2018, 2019, 2021; Trevisin et al., 2022) thus measures the growth rate of the fastest-growing perturbation to an asymptotically stable DFE (or, by extension, to a trajectory slowly departing from it).

Although the Euclidean 𝓁^2^-norm was originally used to measure transient amplification in discrete-time systems (Caswell and Neubert, 2005), as also typically done in reactivity analysis of continuous-time applications (Neubert and Caswell, 1997; Mari et al., 2017), other norms can be used as well. In particular, for epidemiological and disease ecology applications, the 𝓁^1^-norm might be a more appropriate choice, especially for trajectories departing from the DFE, for which only positive perturbations of the infection subsystem are admissible. The 𝓁^1^-norm measures the maximum transient growth of a perturbation based on the absolute deviation (sometimes referred to as taxicab, or Manhattan, distance) from the DFE. As such, it has a more straightforward biological interpretation (i.e., 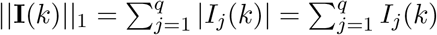 is the total prevalence of infection at time *k*), as compared to the 𝓁^2^-norm, which is instead based on Euclidean distances. For the same reason, the use of the 𝓁^1^-norm has also been advocated in ecological applications, such as the study of animal and plant population dynamics (Townley and Hodgson, 2008; Stott et al., 2010, 2011), and more recently, transient patterns in metapopulations (Harrington et al., 2022). On the contrary, examples of application of the 𝓁^1^-norm in epidemiology and disease ecology are still lacking.

We outline here a framework for the analysis of discrete epidemicity using the 𝓁^1^-norm. All the results presented in the main text refer to this choice. Theory and results concerning the commonly adopted 𝓁^2^-norm are instead reported in SI (section S3). The maximum one-step amplifications evaluated with different norms are expected to be quantitatively different, reinforcing the notion that in reactivity/epidemicity analysis it is always crucial to state which norm is used to evaluate transient amplification (Neubert and Caswell, 1997; Mari et al., 2017; Lutscher and Wang, 2020). The numerical evaluation of both norms applied to matrices is quite straightforward: the 𝓁^1^-norm can be computed as the maximum absolute column sum of the projection matrix **L**(*k*), while the 𝓁^2^-norm can be obtained via singular value decomposition of **L**(*k*) (Horn and Johnson, 2012).

### Discrete epidemicity index

We define our main discrete epidemicity index *ε* as the maximum growth, evaluated in the 𝓁^1^-norm, achieved in one step by any perturbation of the current state of the system. For the sake of simplicity, we assume that the depletion of the susceptible pool is negligible (i.e., we are in a neighborhood of the DFE) and that the control effort does not change over time, namely **L**(*k*) ≡ **L**. Therefore, we have

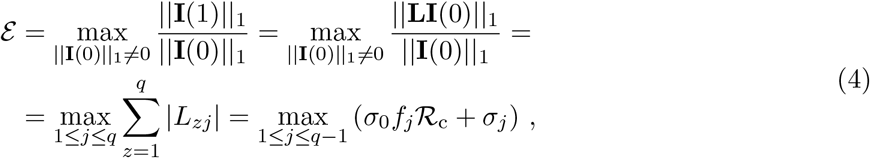

where *L*_*zj*_ is the element at the *z*-th row and *j*-th column of the projection matrix **L**; note that the rightmost-hand side of (4) is obtained in the limit *f*_*q*_ → 0. The sufficient condition to avoid transient outbreaks, which also guarantees the non-recurrence of subthreshold epidemics, is thus *ε* < 1. The maximum one-step growth is achieved by a perturbation that is null for every *j* ∈ {1, …, *q*} except for *j* = argmax_*j*_ (*σ*_0_*f*_*j*_ *ℛ*_c_ + *σ*_*j*_), that is, for the age corresponding to the maximum column sum of the projection matrix (Harrington et al., 2022). This result has an intuitive epidemiological (or ecological) interpretation, which supports again the choice of the 𝓁^1^-norm for this kind of problem: the most critical perturbation is associated with the age (of infection) class that can potentially contribute the most to the (infected) population in the next timestep in terms of the combined effects of recruitment (secondary infections) and survival (permanence within the infected compartment).

Clearly, in the absence of containment measures, the control RN should be replaced with the basic RN. By contrast, in time-variant applications, the discrete epidemicity index needs to be updated with the same frequency as the effective RN, namely *ε*_*k*_ = max_1≤*j*≤*q*−1_ (*σ*_0_*f*_*j*_ *ℛ*_*k*_ + *σ*_*j*_). In this condition, it is thus possible that the critical age of infection changes over time.

### RN epidemicity threshold

Using the discrete epidemicity index defined by (4), it is possible to assess the RN value needed to prevent short-term outbreaks. For the sake of simplicity, we assume that the rate of exit from the infected compartment (because of recovery or death) does not vary much with respect to the age of infection (*γ*(*τ*) ≈ *γ* for all *τ* ‘s), so that we can set *σ*_0_ = exp(−*γ/*2) and *σ*_*j*_ = exp(−*γ*) = *σ* for *j* ∈ {1, …, *q* − 1}, again using a daily step and evaluating survival at discretization midpoints. Imposing *ε* = 1 in (4), we obtain

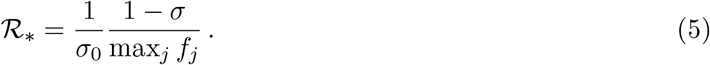

Thus, the condition that must hold to prevent any short-term epidemic outbreak from occurring is RN < *ℛ*_∗_. Note that 0 < *ℛ*_∗_ ≤ 1. In particular, to show that *ℛ*_∗_ ≤ 1, one can observe that, if *σ*_*j*_ = *σ* for *j* ∈ {1, …, *q*}, the fundamental property 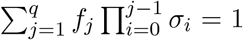 becomes 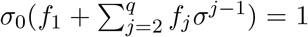. Since 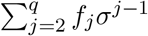 cannot be larger than max 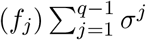, which can be approximated as max_*j*_(*f*_*j*_)[*σ/*(1 − *σ*)] in the limit of large *q*, one obtains *σ*_0_{*f*_1_ + max_*j*_(*f*_*j*_)[*σ/*(1 − *σ*)]} ≥ 1. With straightforward manipulations, it is then possible to conclude that *ℛ*_∗_ ≤ (1 − *σ*)*f*_1_*/*[max_*j*_(*f*_*j*_)] + *σ* ≤ 1.

In general, if *σ*_*j*_ can take different values for different age-of-infection classes, the threshold value *ℛ*_∗_ can be found in the following way. Let *ℛ*_∗,*j*_ be the solution of *σ*_0_*f*_*j*_ *ℛ*_c_ + *σ*_*j*_ = 1, that is, *ℛ*_∗,*j*_ = (1 − *σ*_*j*_)*/*(*σ*_0_*f*_*j*_). Then, *ℛ*_∗_ = min_1≤*j*≤*q*−1_ *ℛ*_∗,*j*_.

### Herd epidemicity threshold

If, in addition to the threshold RN evaluated with (5), information is available also on a pathogen’s *ℛ*_0_, the effort to be put in preventive control measures in order to avoid short-term epidemicity can be usefully assessed using a herd epidemicity threshold (HET) that can be defined in analogy with the herd immunity threshold (HIT), a measure of the fraction of the population that needs to be involved in preventive measures in order to halt the long-term transmission of a pathogen (typically, HIT = max(0, 1 − 1*/ℛ*_0_)). Imposing *ℛ*_0_(1 − HET) = *ℛ*_∗_, we get

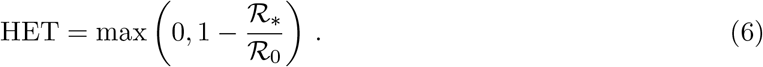

The HETs obtained with (6) can be usefully compared to HIT estimates to identify possible gaps that might exist between the containment effort needed to curb transmission asymptotically vs. in the short term.

### Temporal evolution of transient outbreaks

Transient epidemicity can also be studied on relatively longer timescales than just one step after the perturbation. It is possible to define a so-called amplification envelope (Neubert and Caswell, 1997; Caswell and Neubert, 2005) that quantifies the maximum amplification achieved by any perturbation at a given time *k*, i.e.,

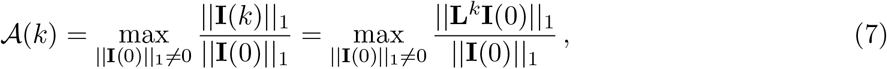

where we have again assumed a negligible depletion of the susceptible pool and time-invariant controls. The amplification envelope at any time *k* ≥ 1 can thus be evaluated as the maximum column sum of matrix **L**^*k*^. The perturbation corresponding to the largest amplification at time *k* is null for all *j* ∈ {1, …, *q*} except for that corresponding to the maximum column sum of **L**^*k*^ (Harrington et al., 2022).

Several features of interest are located along the amplification envelope. According to definition (7), *𝒜*(1) ≡ *ε*. Another point of interest is the peak *𝒜*_max_ = max_*k*_ *𝒜* (*k*), which defines the maximum amplification achieved by any perturbation at any time (say, at *k* = *k*_max_) as measured by the 𝓁^1^-norm (Neubert and Caswell, 1997; Caswell and Neubert, 2005). The inequality *𝒜*_max_ > 1 may define an alternative (and more inclusive) epidemicity condition—albeit one that is less prone to analytical treatment. Finally, if it exists, the first value of *k* > 1 (say, *k* = *k*_end_) for which *𝒜* (*k*) = 1 is also of some relevance because it signals the end of the longest outbreak, defined as the point in time when the maximum amplification becomes smaller than the initial perturbation in the chosen norm.

Note that all the features of the amplification envelope for *k* > 1 are obtained by repeated applications of the projection matrix: as such, all results referring to *k* ≫ 1 have to be considered as upper limits, as they do not account for the nonlinearities of pathogen transmission unless the system is made time-variant and updated estimates of the effective RN are produced over time. In this case, the propagation matrix **L**^*k*^ must be replaced by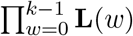.

### Discrete epidemicity in the continuous-time limit

To evaluate the impact of discretization on the results of epidemicity analysis, consider a generic timestep Δ. Intuitively, for small values of Δ, the amplification of any possible perturbation of the DFE should be negligible (independent of the norm considered). More formally, it is immediate to verify that

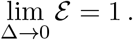

This result, however, may not prevent the amplification envelope from initially having a positive temporal derivative. To show this, we can evaluate the derivative as a difference quotient in the limit of small Δ. Using the first-order approximations *σ*_0_ = exp(−*γ*Δ*/*2) ≈ 1−*γ*Δ*/*2, *σ*_*j*_ = exp(−*γ*Δ) ≈ 1−*γ*Δ for *j* ∈ {1, …, *q* − 1}, and *f*_*j*_ ≈ *ϕ*(*j* − Δ*/*2)Δ, we get

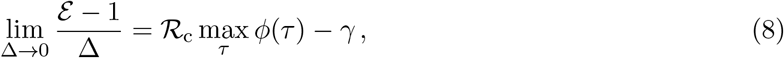

where

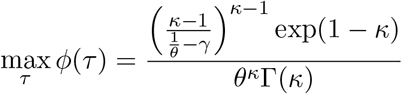

and Γ represents the gamma function. Therefore, to prevent perturbations to an asymptotically stable DFE from initially being amplified, one must impose *ℛ*_c_ to be smaller than

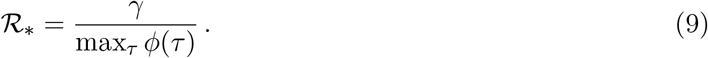

Equivalently, the fraction of the population that must be involved in preventive containment measures must be larger than

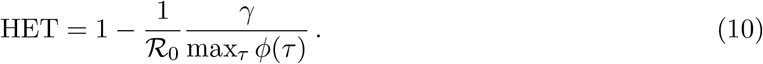

The case Δ ≫ 0 is instead analyzed in SI (section S2).

## Results

Based on the discrete-time formulation of reactivity (Caswell and Neubert, 2005), we have developed a theory of discrete epidemicity (Materials and Methods) and showed that recurrent transient epidemics can be avoided if a discrete epidemicity index *ε*, corresponding to the maximum one-step amplification of any perturbation to the system state, is < 1. As stated above, assessing the growth of epidemiological perturbations over time requires the definition of a suitable norm (Neubert and Caswell, 1997) or output transformation (Mari et al., 2017): in this section, which is devoted to applying the method to well-known respiratory infections, we use the 𝓁^1^-norm, basically corresponding to total infection prevalence (i.e., computed over all ages of infection). If we use this norm, *ε* simply corresponds to the largest one-step-ahead contribution by any single age-of-infection class to disease prevalence within the population, considering both new infections and permanence within the infected compartment.

As an example, using the parameter estimates for ancestral SARS-CoV-2 (Table 1) and a value of *ℛ*_c_ = 0.95 (which could represent a minimum target for control interventions aimed at mere herd immunity), one gets *ε* = 1.24 (corresponding to a 24% case increase in one day). Therefore, although long-term transmission cannot be established under such conditions, transient epidemic outbreaks are still possible (Figure 2). Imposing *ε* = 1, the threshold value of the RN (*ℛ*_∗_) and, equivalently, the herd epidemicity threshold (HET, corresponding to the share of the population to be involved in containment actions in order to prevent transient epidemic outbreaks) can be analytically computed. For COVID-19, one would get *ℛ*_∗_ = 0.24 (≪ 1) and HET = 0.93 (≫ HIT = 0.72). Numerically, it is also possible to evaluate the amplification envelope *𝒜* (*k*), that is the maximum amplification attainable by any isolated perturbation after a given time *k*, and extract some important reactivity metrics (in addition to *ε*, which by definition coincides with *𝒜* (1)), namely: the maximum amplification at any time (*𝒜*_max_, which turns out to be 2.0 for COVID-19, corresponding to a 100% increase); the time to reach the maximum case surge (*k*_max_, 22 days in the case of COVID-19); and the duration of the longest outbreak (*k*_end_, 114 days for COVID-19; see again Figure 2, in which *ℛ*_c_ = 0.95). A complete example of epidemicity analysis applied to another disease (measles) is reported in SI (section S2 and Figures S1–S2; in the latter figure, in particular, sensitivity analysis also shows that a daily discretization time step is a reasonable trade-off choice between feasibility and accuracy).

**Figure 2:**
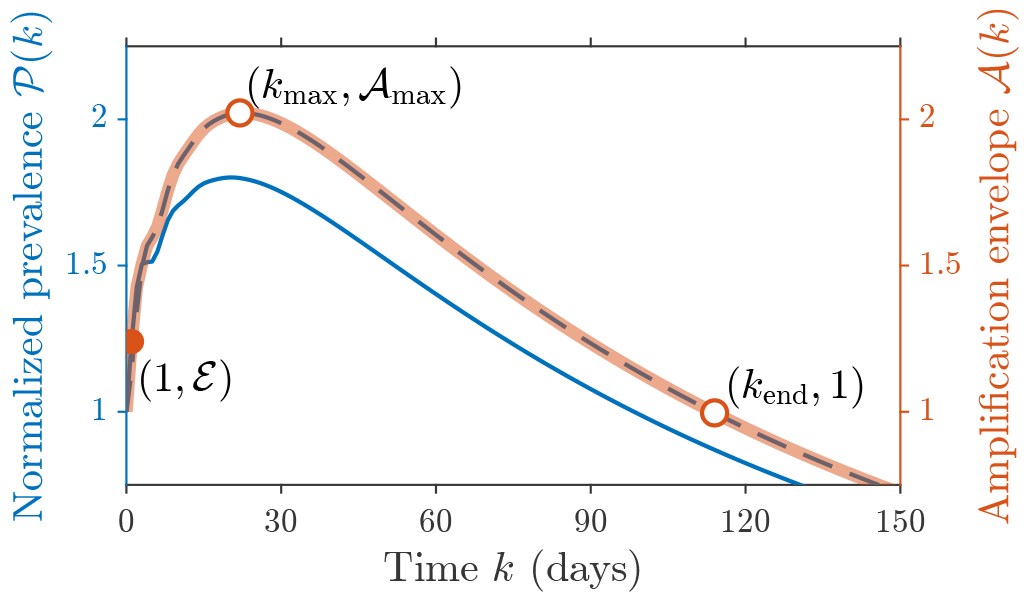
Transient epidemicity analysis for COVID-19 for a subthreshold value of the control RN. Two examples of model simulations, corresponding to the perturbations of the system state characterized by maximum amplification in one step (solid blue line, left axis) or overall (dashed blue line, left axis; note that *𝒫* (*k*) = ||**I**(*k*)||_1_*/*||**I**(0)||_1_, see Materials and Methods), and the amplification envelope (red line, right axis). The three dots (coordinates in parentheses) mark the one-step maximum amplification, the maximum amplification overall, and the point in time when the amplification envelope returns below the initial perturbation size; some of these features can be computed analytically, while others require to numerically evaluate the envelope (color-filled vs. white-filled dots). Parameter values: AV = 5.2 days, SD = 1.7 days, 1*/γ* = 11.7 days (Table 1), *ℛ*_c_ = 0.95. In all analyses, the depletion of the susceptible pool is considered negligible and the RN is kept constant over time.

Figure 3 reports summary results not only for COVID-19 and measles but also for 13 other respiratory viruses (Table 1). By using again a value of *ℛ*_c_ = 0.95, the *ε* values range between 1.08 (MERS) and 1.43 (Influenza A(H3N2)), with a median value of 1.23, while *𝒜*_max_ can reach between 1.44 (MERS) and 2.30 (rubella), with a median of 1.79 (Figure 3(a)). Regarding temporal dynamics (Figure 3(b)), the median *k*_max_ is 22 days, while the median *k*_end_ is 117 days, with the shortest or longest outbreaks expected for Influenza A(H1N1)pdm09 or mumps, respectively (in terms of both *k*_max_ and *k*_end_). The *ℛ*_∗_ values vary between 0.10 (rubella) and 0.51 (MERS), with a median value of 0.26 (Figure 3(c)). When considering also basic RNs, HITs range between 0 (MERS, for which *ℛ*_0_ < 1) and 0.93 (measles), with a median value of 0.72; by contrast, HETs range between 0.28 (MERS) and 0.99 (measles), with a median value of 0.93 (Figure 3(d)). The widest gaps between HITs and HETs are recorded for pathogens with the lowest basic RN values, namely those responsible for Influenza A(H1N1)pdm09, Influenza A(H3N2), Influenza B, MERS, and RSV, all of which are characterized by HET-to-HIT ratios > 3. A sensitivity analysis of these results (SI, Section S2 and Figures S3–S8) reveals remarkable robustness to changes in the choice of the underlying parameters for all metrics related to one-step perturbation amplification (*ε, ℛ*_∗_, and HET), as well as for the maximum amplitude achieved by any perturbation at any time (*𝒜*_max_), while the metrics associated with the temporal characteristics of the amplification envelope may be characterized by comparatively larger variability ranges (especially *k*_max_). Switching to the 𝓁^2^-norm to evaluate transient amplifications (SI, section S3 and Figure S9) does not alter qualitatively the results, but produces quantitative differences that typically yield an underestimation of the control effort needed to prevent transient epidemicity. Therefore, the 𝓁^1^-norm not only yields results that are more amenable to biological interpretation than those obtained with the 𝓁^2^-norm but also provides a more conservative approach to the design of containment measures.

**Figure 3:**
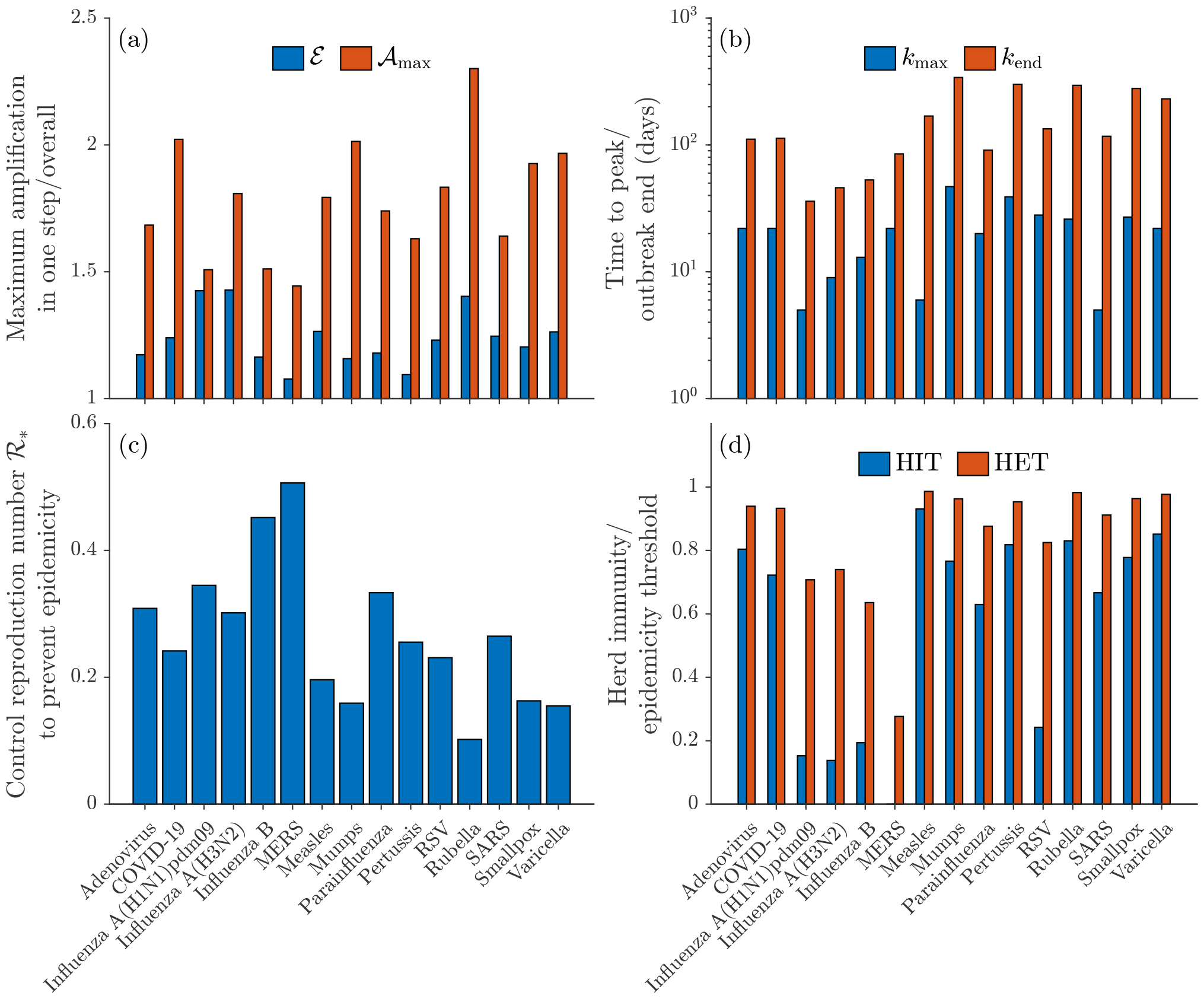
Summary of the transient epidemicity results for different respiratory viruses. (a) Maximum amplification achievable in one step (blue) and overall (red), assuming *ℛ*_c_ = 0.95. (b) Time to maximum prevalence peak (blue) and maximum outbreak duration (red), using the same *ℛ*_c_ value as in panel (a). (c) Threshold RN to prevent transient epidemic outbreaks. (d) Fraction of the population to be involved in preventive control actions to avoid long-term pathogen circulation (blue) or short-term epidemicity (red); note that the blue bar is missing for MERS because *ℛ*_0_ < 1 for the causative pathogen. Parameter values as in Table 1.

Thanks to to its modest data requirements, short-term epidemicity analysis can also be applied to emerging pathogens (Morens et al., 2004; Morens and Fauci, 2020; Saad-Roy et al., 2020), as long as some basic knowledge of the relevant transmission timescales is available. To show the prognostic value of our approach, we consider a large set of realistic parameterizations of the transmission process (in terms of generation time distribution, infection duration, and basic RN) that are inspired by the parameter ranges reported in Table 1—but not necessarily linked to a specific, known pathogen— and evaluate the value of HET that guarantees the prevention of short-term epidemic outbreaks (Figure 4). Our results show that short-term epidemicity is typically harder to prevent (larger HET) for pathogens with longer average generation time (AV), smaller standard deviation of the generation time distribution (SD), longer infection duration (1*/γ*, with *γ* being the removal rate from the infected compartment), and higher baseline transmissibility (epitomized by *ℛ*_0_). In other words, prolonged infections (low *γ*, coupled with long AV) with a marked, age-of-infection-dependent, infectivity peak (high *ℛ*_0_ and small SD) are those for which the risk of transient, possibly recurrent outbreaks is highest, consistent with the proposed expression for HET (Materials and Methods). In general, avoiding short-term epidemicity may require participation in preventive interventions (typically vaccination) of very large shares of the exposed population, with HET exceeding 0.95 for many realistic parameterizations.

**Figure 4:**
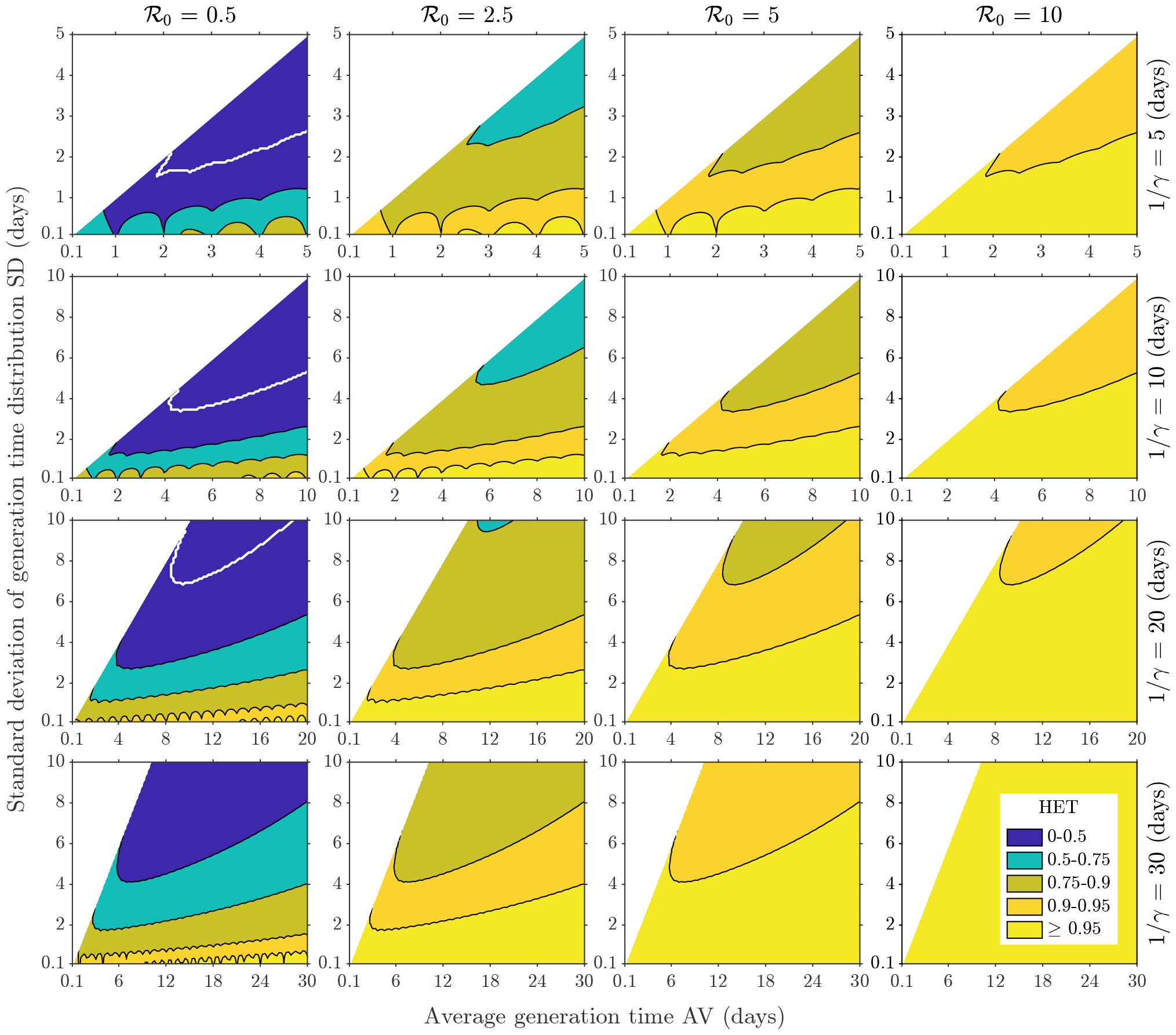
Herd epidemicity threshold to prevent short-term epidemic outbreaks in emerging pathogens with given characteristics. The value of HET (color coded) is calculated for different parameterizations of the generation time distribution (in terms of AV and SD), duration of the infection (1*/γ*, rows), and basic RN (*ℛ*_0_, columns), which have been generated consistently with the ranges reported in Table 1. The white curves separate the parameter combinations for which HET = 0 (because *ℛ*_0_ < *ℛ*_∗_ < 1, above each curve) from those with HET > 0 (below each curve). The parameter combinations for which HET has not been evaluated are those leading to generation time distributions with null mode (shown in white). Note that the jagged profiles of the contour curves at low values of SD are related to the choice of a daily discretization timestep.

## Discussion

Our main result is the surprising difference generally found between the unit value of the control or effective RNs, commonly the target benchmark of containment policies, and the actual RN thresholds (*ℛ*_∗_) that warrant the achievement of safer conditions (i.e., the avoidance of transient flare-ups), with values falling in a 0.10–0.51 range for the spectrum of viral infections investigated here. Implications for policy-making abound because extended containment measures are needed to prevent transient responses from producing a significant resurgence of infections. Regardless of practical feasibility, full awareness of the consequences of epidemiological interventions seems essential. Similarly, HETs show significantly higher values than the corresponding HITs. An arresting example is indeed MERS, which does not have a positive HIT due to its subthreshold basic RN but has a HET of around 30%. This result may suggest why MERS caused serious outbreaks in several countries (Kucharski and Althaus, 2015) despite its low baseline transmissibility. Similar observations apply also to influenza strains characterized by fairly low basic RNs (Trentini et al., 2022) or measles outbreaks observed despite strong containment efforts likely leading to a subthreshold control RN (Blumberg et al., 2015).

Both *ℛ*_∗_ and HET depend on some basic infectious disease characteristics, as shown in Figure 4. Not only higher basic RN, but also longer average generation time and mean duration of the infectious cycle correlate with higher HET and lower *ℛ*_∗_. Longer generation times extend the infectiousness window and provide momentum for future build-ups regardless of possibly ongoing containment measures. Similarly, increased epidemicity occurs when the mean removal time increases, reinforcing the danger of undetected infections. In contrast, higher standard deviation of the generation time distribution correlates with lower HET and higher *ℛ*_∗_. This result echoes recent findings (Lehtinen et al., 2021) about the relationship between the variance of the generation time distribution and the controllability of an outbreak, with smaller variance leading to lower chances of effective epidemic containment because, e.g., of stricter conditions for successful contact tracing. Moreover, using the HIT as a control target may still yield infection flare-ups developing often and rapidly (if HET > HIT), even when a disease is endowed with relatively low basic RN. This result calls for wide vaccination campaigns against diseases with a limited generation of immunity-resistant strains, possibly combined with other containment measures, to guarantee that not only the HET but also the HIT condition are met, thus securing *ε* < 1.

Our study is not devoid of limitations. One is the minimalist assumption of first-order decay from the pool of infectious cases. Although this choice underpins the ease of computation, it does not conceptually affect our results. Another limitation concerns the possible use of space-explicit renewal equations that quantify infections imported to/exported from each community. Including detailed spatial connectivity, when supported by the kind of mobility data commonly available by tracking mobile phones, would imply a more accurate description of transmission mechanisms (Trevisin et al., 2023). Thus a natural follow-up of this study should pursue the computation of spatially explicit epidemicity. In spatially connected communities, infections generated in one place can foster pathogen spread via case seeding and transient responses that may coalesce, in both space and time, into large surges in the number of infections (Mari et al., 2019, 2021). Similarly, non-spatial forms of clustering within the population (as determined, for instance, by differential compliance with the implementation of containment measures; Salathé and Bonhoeffer, 2008) could produce similar transient dynamics and would perhaps be worth exploring. Another aspect that deserves attention is the impact of the evolution of variants (Koelle et al., 2022). Many diseases are characterized by the coexistence of several strains of the same pathogen in a given population. They might, and in many cases do, have different epidemiological parameters and thus different epidemicity thresholds in terms of both RN epidemicity threshold and HET. Therefore, future developments of our work should consider a more accurate model that includes this additional source of heterogeneity.

Three remarks support the relevance of the sufficient RN approach pursued here. The first concerns the limits of our capability to track infection seeding regardless of the geographic domain of interest (whether it be a regional context, a whole country, or even a continent), which should warrant a precautionary approach. The second is related to the overall relevance of spatial effects in estimating RNs and their related HITs (and, possibly, HETs). It has been shown that notable differences between space-implicit and explicit RNs arise only when marked heterogeneity of connected communities affects the demographic and epidemiological attributes involved (Gatto et al., 2020; Trevisin et al., 2023). Thus, we expect our approach, even if spatially implicit at present, to capture the essence of the differences between the necessary and sufficient conditions for epidemicity in most cases. The third and final remark concerns the relative simplicity of the techniques used in this work, which may be applied straightforwardly not only to other respiratory viral diseases, but also to quite different types of infection. Considered together, these considerations provide a clear indication of the possible usefulness of including epidemicity metrics to complement the toolbox of quantitative epidemiologists and disease ecologists.

## Data Availability

This work has no associated primary data. The secondary data used to parameterize the different viral respiratory infections are described in the Supporting Information.

## Acknowledgments

L.M. acknowledges funding from the Italian Ministry of University and Research through via the project “Epidemiological data assimilation and optimal control for short-term forecasting and emergency management of COVID-19 in Italy” (FISR 2020IP 04249). C.T. and A.R. acknowledge funding from the Swiss National Science Foundation via the project “Optimal control of intervention strategies for waterborne disease epidemics” (200021-172578). A.R. acknowledges funding from Fondazione Cassa di Risparmio di Padova e Rovigo (Italy) through its grant 55722. The authors wish to thank Enrico Bertuzzo, Renato Casagrandi, Stefano Miccoli, and Damiano Pasetto for useful discussions. The authors declare no conflicts of interest.

## Author contributions

L.M. and M.G. conceived the ideas and developed the theoretical framework, with substantial contributions by C.T. and A.R.; L.M. collected the data; L.M. and C.T. performed the numerical analyses; all authors participated in the interpretation of the results; L.M. and M.G. led the writing of the manuscript; all authors substantially contributed to the drafts and gave final approval for publication.

## Data and code availability

This work has no associated primary data. The secondary data used to parameterize the different viral respiratory infections are described in the Supporting Information. The main code to perform discrete epidemicity analysis will be made available at https://github.com/lorenzo-mari/discrete-epidemicity.

