## Supplementary Information for "Sufficient reproduction numbers to prevent recurrent epidemics"

#### Supporting Information

Lorenzo Mari[a,1], Cristiano Trevisin[b], Andrea Rinaldo[b,c], Marino Gatto[a,2]

[a]Dipartimento di Elettronica, Informazione e Bioingegneria, Politecnico di Milano, Via Ponzio 34/5, Milano 20133, Italy

[b]Laboratory of Ecohydrology, École Polytechnique Fédérale de Lausanne, Station 2, Lausanne 1015, Switzerland

[c]Dipartimento di Ingegneria Civile, Edile e Ambientale, Università di Padova, Via Marzolo 9, Padova 35131, Italy

#### S1 Parameterization of selected viral respiratory infections

To be applied, the SIR model with age-of-infection structure described in the Materials and Methods section of the main text (equation (1)) and its discrete counterpart (equation (2)) require only modest data inputs, basically consisting of the distribution of generation times, the average duration of the infection, and an estimate of the basic reproduction number (RN). In the following, we report the details of the parameterization of the model for each of the 15 viral respiratory infections considered in the main text. Table 1 in the main text provides a summary of the resulting parameter values.

Note that, whenever the distribution of generation times is not readily available, we use instead the distribution of serial intervals, defined as the time intervals between successive cases in a transmission chain, measured at the onset of clinical symptoms, despite the fact that the two distributions may not coincide for all pathogens (Fine, 2003; Lehtinen et al., 2021). We always assume that generation times/serial intervals follow a gamma distribution with shape parameter $\kappa > 1$ (so that the distribution mode is positive) and scale parameter $\theta > 0$. These two parameters are related to the average value (AV) and standard deviation (SD) of the distribution according to $\kappa = \mathrm{AV}^2/\mathrm{SD}^2$ and $\theta = \mathrm{SD}^2/\mathrm{AV}$ (or, equivalently, $\mathrm{AV} = \kappa\theta$ and $\mathrm{SD}^2 = \kappa\theta^2$).

#### Adenovirus infection

We use the serial interval distributions estimated in ref. (Wang et al., 2021) for low altitudes, yielding AV = 7.1 days and SD = 2.6 days. As for the duration of the infection, we use the temporal progression proposed in ref. (Zhang et al., 2022), where the authors estimated an incubation time of 4 days (average of three viral types) and a different recovery period for symptomatic (34% of total cases on average) vs. asymptomatic infections (9.0 days vs. 6.7 days on average), with a weighted average of 7.5 days, for a total average duration of the infected period of 11.5 days (hence, $\gamma = 1/(11.5\,\text{days}) = 0.087\,\text{days}^{-1}$). The estimate of the basic RN ($\mathcal{R}_0 = 5.1$) is taken from ref. (Guo et al., 2020).

#### COVID-19 (SARS-CoV-2 infection)

We rely on the generation time distribution estimated in ref. (Ganyani et al., 2020), where the values AV = 5.2 days and SD = 1.7 days were reported. To quantify infection duration, we use the parameters calibrated in ref. (Gatto et al., 2020), which proposed an incubation period of 3.3 days, a pre-symptomatic phase of 0.75 days, and a permanence within the infected compartment that depends upon the intensity of symptoms, namely 7.2 days for asymptomatic carriers (estimated to be 75% of all cases) and 9 days for symptomatic infections (also considering disease-related mortality), with a weighted average of 7.6 days; combining all these factors we get an average duration of infection of 11.7 days, corresponding to $\gamma = 0.086\,\text{days}^{-1}$. The same authors also estimated $\mathcal{R}_0 = 3.6$ for the original viral strain.

#### Influenza A(H1N1)pdm09 infection

For this pandemic strain of influenza, we use the serial interval distribution data published in a systematic review on respiratory infections (Vink et al., 2014), namely AV = 2.8 days and SD = 1.0 days. For infection duration, we use the timeline published in ref. (Wang et al., 2014), where an incubation period of 1.1 days and an infectious period of 2.5 days were proposed, for a total duration of the infection of 3.6 days (hence, $\gamma = 0.28\,\text{days}^{-1}$). For the basic RN, we use the average value $\mathcal{R}_0 = 1.18$ found in ref. (Trentini et al., 2022).

#### Influenza A(H3N2) infection

We use the same source (Vink et al., 2014) as for the previous influenza virus strain to characterize the distribution of serial intervals, i.e., AV = 2.3 days and SD = 0.80 days. For the duration of disease, we use the central estimate of 4.5 days proposed in ref. (Casagrandi et al., 2006), from which we get

$\gamma = 0.22\,\text{days}^{-1}$, while we rely on the average estimate of $\mathcal{R}_0 = 1.16$ proposed in ref. (Trentini et al., 2022) for the basic RN.

#### Influenza B infection

For the serial interval distribution, we use the estimates of AV = 3.7 days and SD = 2.0 days proposed in ref. (Levy et al., 2013). For the length of the infected period, we use the positive virus isolation results in ref. (Sato et al., 2005), where the authors reported a duration of 6.2 days for untreated patients, hence $\gamma = 0.16\,\text{days}^{-1}$. For the basic RN, we use the average value $\mathcal{R}_0 = 1.24$ found in ref. (Trentini et al., 2022).

#### Middle East respiratory syndrome (MERS)

For this human coronavirus infection, we rely on the serial interval information provided in ref. (Cauchemez et al., 2016), namely AV = 6.8 days and SD = 4.1 days. For the duration of disease, we use the estimates from South Korea provided in ref. (Rui et al., 2022), in which the authors proposed an incubation period of 6.9 days and duration of the infectious period of 3.9 days (averaged over asymptomatic infections, which represent 7% of the total, and symptomatic ones—considering also disease-related mortality for the latter), from which we get a total infection duration of 10.8 days (hence, $\gamma = 0.093\,\text{days}^{-1}$). As for the basic RN, we use the early estimate of $\mathcal{R}_0 = 0.7$ reported in ref. (Kucharski and Althaus, 2015).

#### Measles

We use the serial interval distribution data compiled in the systematic review reported in ref. (Vink et al., 2014), that is, AV = 11.7 days and SD = 2.5 days. We consider 14 days as the duration of the infection, as suggested in ref. (Bjørnstad et al., 2002), from which we obtain $\gamma = 0.071\,\text{days}^{-1}$, and the estimate of $\mathcal{R}_0 = 14.5$ proposed by ref. (van den Driessche, 2017).

#### Mumps

We use again the serial interval distribution data published in ref. (Vink et al., 2014), where the values AV = 18.0 days and SD = 3.5 days were proposed. As for the duration of disease and the basic RN, we use the estimates available from ref. (Li et al., 2018), where the authors reported an incubation period of 19 days and a 12 day-long permanence within the infected compartment (central values), for a total infection duration of 31 days (from which we get $\gamma = 0.032\,\text{days}^{-1}$). From the same source, we also get an estimate of $\mathcal{R}_0 = 4.28$.

#### Parainfluenza

As for the serial interval distribution, we use the data reported in ref. (Banatvala et al., 1964), namely AV = 5.2 days and SD = 2.2 days. As for the duration of disease and the basic RN, we use the estimates proposed in ref. (Reis and Shaman, 2018), where the authors reported an average duration of illness of 9.8 days (hence, $\gamma = 0.10\,\mathrm{days}^{-1}$) and a value of $\mathcal{R}_0 = 2.7$.

#### Pertussis

We use the serial interval distribution data compiled in ref. (Vink et al., 2014), namely AV = 22.8 days and SD = 6.4 days. The numerical values of the duration of disease (28 days, including an incubation period of about 7 days, from which $\gamma = 0.036\,\mathrm{days}^{-1}$) and the basic RN $\mathcal{R}_0 = 5.5$ are taken from ref. (Hethcote, 1997).

#### Respiratory Syncytial Virus (RSV) infection

We use again the serial interval distribution data published in ref. (Vink et al., 2014), where the values AV = 7.5 days and SD = 2.1 days were proposed. As for the duration of illness, we rely on the information provided in ref. (Hogan et al., 2016), where a latent period of 4 days and an infectious period of 9 days were reported, for a total duration of the infection of 13 days (hence, $\gamma = 0.077\,\mathrm{days}^{-1}$). The value of the basic RN $\mathcal{R}_0 = 1.32$ is obtained from the same literature source.

#### Rubella

We use again the serial interval distribution data available from ref. (Vink et al., 2014), that is, AV = 18.3 days and SD = 2.0 days. For the duration of disease, we consider the average value of 20.1 days proposed in ref. (Thompson, 2016) while considering models accounting for the whole course of the infection, from which we get $\gamma = 0.050\,\mathrm{days}^{-1}$. From the same source, we also take the value of the basic RN $\mathcal{R}_0 = 5.9$ (average value).

#### Severe acute respiratory syndrome (SARS)

We consider estimates of the serial interval distribution provided by a classic study on SARS transmission (Lipsitch et al., 2003), where the authors found AV = 10.0 days and SD = 2.8 days. From the same study, we also infer the typical timescale of infection progression, with incubation and infectious periods approximately lasting 5 days each (the latter, also including the effect of disease-related mortality), from which we obtain $\gamma = 0.10\,\mathrm{days}^{-1}$, as well as the basic RN $\mathcal{R}_0 = 3.0$.

##### Smallpox

We use the serial interval distribution data published in ref. (Vink et al., 2014), namely AV = 17.6 days and SD = 3.2 days. As for the duration of the infection, we use the temporal progression proposed in ref. (Gani and Leach, 2001), where a latent period of 14.6 days and an infection duration of 8.6 days (also accounting for disease-related mortality) were reported, for a total duration of the infected period of 23.2 days (hence, $\gamma = 0.043\,\text{days}^{-1}$). For the basic RN, we use the estimate of $\mathcal{R}_0 = 4.5$ proposed in ref. (van den Driessche, 2017).

##### Varicella (chickenpox)

We resort once again to the data from the systematic review described in ref. (Vink et al., 2014), from which we have AV = 14.0 days and SD = 2.4 days. For the duration of infection, we refer to ref. (Garnett and Grenfell, 1992), where the authors considered an incubation period of 14 days followed by an infectious period of 5 days; combining these two periods, we get an estimate of infection duration of 19 days, from which we get $\gamma = 0.053\,\text{days}^{-1}$. The estimate of the basic RN is instead taken from ref. (Tang et al., 2017), from which we get $\mathcal{R}_0 = 6.73$.

#### S2 Supporting results

In this section, we present some results complementing the principal findings described in the main text. All the results reported here are obtained with the same $\ell^1$-norm used in the main text.

##### Transient epidemicity analysis for a prototypical viral disease: the case of measles

We perform a complete epidemicity analysis for one of the viral diseases introduced above and analyzed in the main text. We specifically select measles for its "average" characteristics in terms of the transmission timescales presented in Table 1 in the main text (AV = 11.7 days vs. an average of 10.2 days evaluated across infections; SD = 2.5 days vs. an average of 2.6 days; $1/\gamma = 14$ days vs. an average of 14.4 days); by contrast, we are less concerned about its uncharacteristically high basic RNs ($\mathcal{R}_0 = 14.5$ vs. an average of 4.1) because we mostly focus on controlled situations in which $\mathcal{R}_c$ should be below the unit threshold.

For measles, if we use the value $\mathcal{R}_c = 0.9$, we find $\mathcal{E} = 1.25$, corresponding to a 25% maximum prevalence increase in one day. For this value of $\mathcal{R}_c$, any outbreak is still eventually destined to fade; however, transient epidemic waves are still possible, as shown in Figure S1. Specifically, in panel (a), we show two numerical simulations of model (2) corresponding to the initial conditions leading to the maximum one-step growth (solid blue line) and overall (dashed blue line), as well as the amplification envelope (red line). The maximum amplification of any perturbation to the DFE occurs after $k_{\max} = 6$ days and corresponds to a 73% prevalence increase with respect to the initial perturbation size ($\mathcal{A}_{\max} = 1.73$). It may take almost three months ($k_{\mathrm{end}} = 86$ days) for disease prevalence to fall below the level of the initial perturbation, suggesting that transient pathogen transmission dynamics can have relatively long-lasting epidemiological implications. An interesting feature of the simulations reported here is the presence of damped oscillations towards the asymptotically stable DFE: these prevalence fluctuations are linked to the presence of (possibly many) complex conjugate sub-dominant eigenvalues in the spectrum of the Leslie projection matrix (Caswell, 2000). To avoid transient epidemicity, the control RN should be below $\mathcal{R}_* = 0.20$, a condition that in this case prevents the amplification of any perturbation to the DFE both over the first timestep and at any later time (panel (b)). This value of the control RN corresponds to a HET of 0.99, a figure that has to be compared to a HIT value of 0.93 (i.e., 99% of the population needs to be involved in preventive control measures in order to avert transient outbreaks, while the involvement of 93% of the population is sufficient to halt disease spread asymptotically). Finally, panel (c) shows some temporal features of the amplification envelope for different values of the control RN, with numerical results confirming the intuitive expectation that longer outbreaks should be observed for values of $\mathcal{R}_c$ approaching the unit threshold.

#### Sensitivity analysis: temporal resolution

In the analysis of the measles case study just described (and in all other analyses reported in the main text), we applied a daily discretization timestep. Before proceeding further, it is this important to establish whether this choice may produce any relevant impacts on the results of short-term epidemicity analysis. To this end, we resort again to the prototypical example of measles.

Let us thus consider a generic discretization timestep $\Delta$. The limit case $\Delta \to 0$ has already been considered analytically in the Materials and Methods section of the main text. The case $\Delta \gg 0$ must instead be studied numerically. Some sensitivity analysis results with respect to the choice of $\Delta$ for the case study of measles are shown in Figure S2. The computation of the amplification envelope is indeed quite heavily influenced by the discretization timestep; however, the choice of $\Delta = 1$ day still seems to produce a reasonable approximation of the process when compared to shorter (and likely more difficult to apply in practice) timesteps (panel (a)). In terms of one-step epidemicity, the values of $\mathcal{E}$ tend progressively to one as $\Delta$ is decreased from one day to shorter values, while the derivative of the amplification envelope at the time of the perturbation converges to the theoretical value evaluated with (8) (Materials and Methods, main text); on the other hand, the evaluation of $\mathcal{E}$

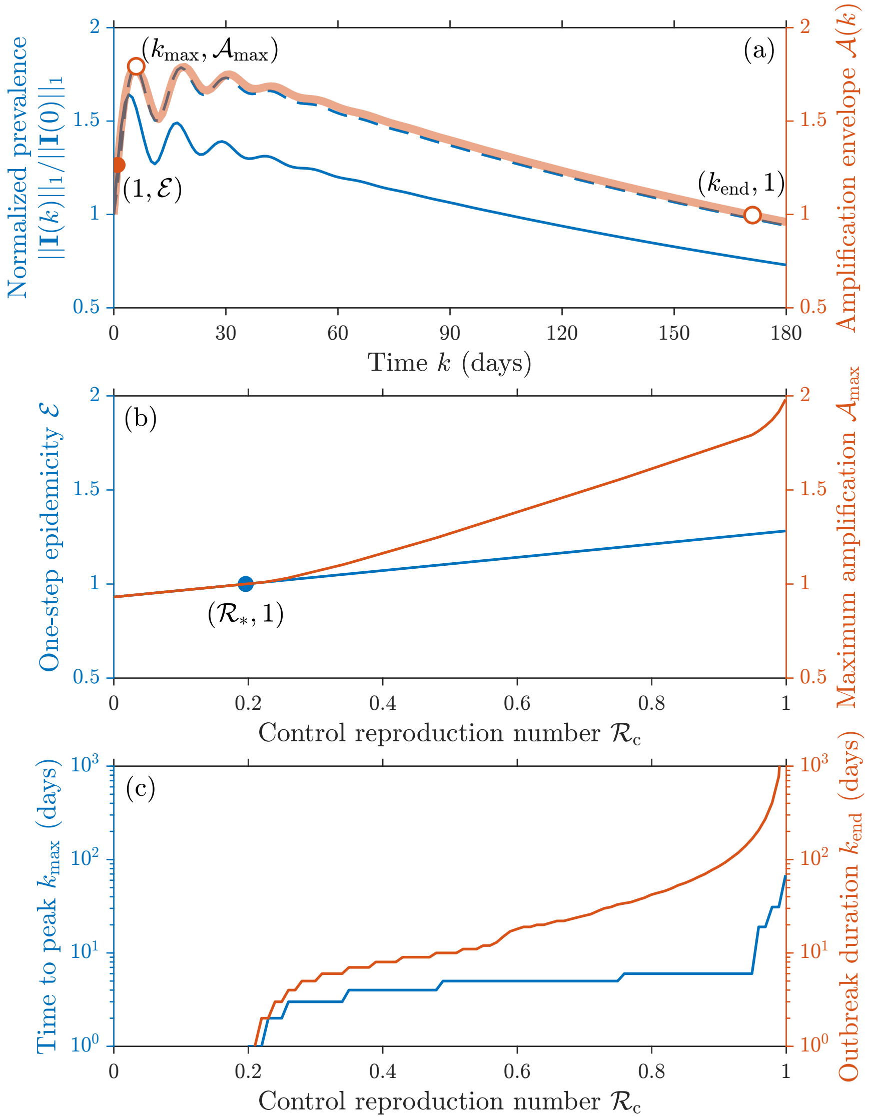


Figure S1: Transient epidemicity analysis for measles. (a) Two examples of model simulations, corresponding to the perturbations of the DFE characterized by maximum amplification in one step (solid blue line, left axis) or overall (dashed blue line, left axis), and the amplification envelope obtained for $\mathcal{R}_c = 0.95$ (red line, right axis). The three dots mark the one-step maximum amplification, the maximum amplification overall, and the point in time when the amplification envelope returns below the initial perturbation size; some of these features can be computed analytically, while others require to numerically evaluate the envelope (color-filled vs. white-filled dots). (b) One-step amplification (blue curve, left axis) and maximum amplification overall (red curve, right axis) for different values of $\mathcal{R}_c$. The blue curve, including the threshold value $\mathcal{R}_*$ of the RN to avoid epidemicity, can be evaluated analytically, while the red curve is obtained numerically. (c) Temporal features of transient epidemic outbreaks for different values of the control RN (blue, left axis: number of timesteps needed to reach the peak of the amplification envelope; red, right axis: maximum duration of the outbreak), obtained through numerical simulations of the model. Parameter values: AV = 11.7 days, SD = 2.5 days, $1/\gamma = 14$ days (Table 1 in the main text). See Section S2 for an extended description of the results shown in this figure.

(as well as of the derivative of the amplification envelope) becomes increasingly more uncertain as $\Delta$ is increased from one day to longer values, especially when $\Delta$ approaches the timescale associated with the peak of the amplification envelope (panel (b)). Therefore, the choice of too large of a discretization timestep may lead to missing important features of the amplification envelope over different timescales. Similar remarks apply to the evaluation of the other one-step epidemicity metrics, namely $\mathcal{R}_*$ and HET (panel (c)). In particular, the threshold value of the control RN needed to prevent short-term outbreaks converges to the analytical approximation given in (9) in the limit $\Delta \to 0$ (Materials and Methods, main text); as a result, the herd epidemicity threshold too converges to a definite value that can be evaluated analytically in the same limit approximation with (10) (Materials and Methods, main text). Quite importantly, in terms of the overall robustness of the choice of a daily discretization timestep, the value of $\mathcal{R}_*$ obtained with $\Delta = 1$ day differs by less than 1% from that computed in the theoretical limit $\Delta \to 0$; an even smaller difference, in the order of 0.01%, is found for HET. By contrast, choosing a value of $\Delta \gg 1$ would result in inaccurate estimates of both $\mathcal{R}_*$ and HET.

#### Sensitivity analysis: parameter variations

A sensitivity analysis of the results shown in Figure 3 in the main text with respect to variations of the baseline parameterization for each respiratory virus is shown in Figures S3–S8 for six epidemicity-related metrics, namely: $\mathcal{E}$ (Figure S3), $\mathcal{A}_{\max}$ (Figure S4), $k_{\max}$ (Figure S5), $k_{\text{end}}$ (Figure S6), $\mathcal{R}_*$ (Figure S7), and HET (Figure S8). We find that the metrics related to the maximum one-step amplification ($\mathcal{E}$, $\mathcal{R}_*$, and HET) are typically characterized by smaller sensitivity ranges than the considered parameter variations (the only exception being the HET metric for MERS, the only disease with $\mathcal{R}_0 < 1$ among those considered in Table 1 in the main text), thereby showing considerable robustness to changes in the underlying parameter choice. For these metrics, it is also generally possible to identify consistent directions of change (i.e., the sensitivity values evaluated for all infections share the same sign for a positive/negative variation of the considered parameter), with some possible exceptions for the smallest sensitivity values (e.g., for $\mathcal{E}$). Interestingly, the $\mathcal{A}_{\max}$ metric, which is evaluated at $k = k_{\max} > 1$, is also found to have sensitivity ranges that are generally smaller than the considered parameter variations. This result is even more remarkable noting that the $k_{\max}$ metric is the one showing the largest sensitivity to parameter variations, especially with respect to changes in the average generation time and infection duration. In this case, though, there can be considerable variability in the direction of change of the metric, in particular when evaluated against variations in the average generation time. Finally, the $k_{\text{end}}$ metric seems to be characterized by noteworthy robustness with respect to variations of the parameters associated with the generation

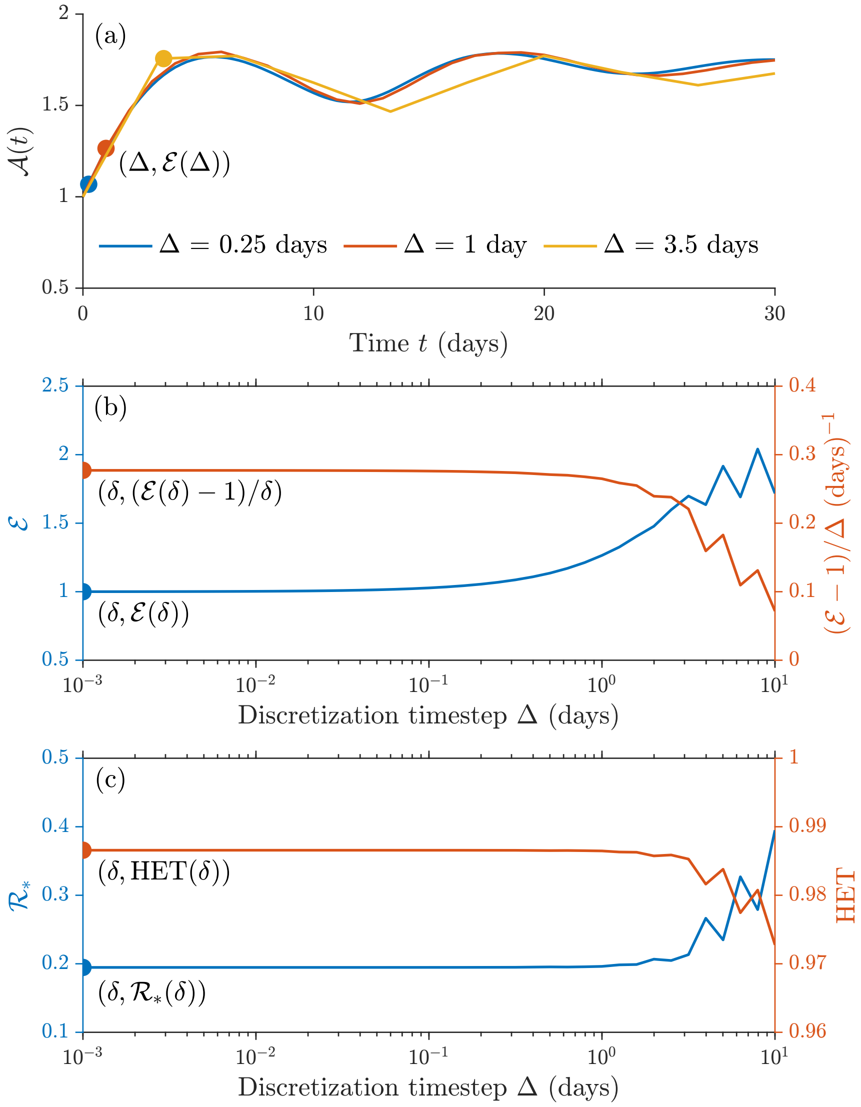


Figure S2: Sensitivity analysis of the epidemicity analysis results for measles with respect to changes of the discretization timestep $\Delta$. (a) Amplification envelopes obtained with three different values of $\Delta$ for $\mathcal{R}_c = 0.95$. One-step maximum amplifications (points marked with filled dots) have been computed analytically after suitably adapting the entries of the Leslie projection matrix to account for the value of $\Delta$. (b) One-step maximum amplification (blue, left axis) and derivative of the amplification envelope at the time of the perturbation (red, right axis) evaluated for different values of $\Delta$. The points marked with filled dots have been computed analytically in the limit $\delta \to 0$ (graphically shown for $\Delta = \delta = 10^{-3}$ days). (c) As in (b), for the threshold value of the control RN to avoid epidemicity (blue, left axis) and the herd epidemicity threshold (red, right axis). Parameter values as in Figure S1. See Section S2 for an extended description of the results shown in this figure.

time distribution, less so with respect to variations of infection duration.

### S3 Epidemicity analysis using the $\ell^2$-norm

So far, we have always used the $\ell^1$-norm to assess the transient growth of perturbations to a stable DFE. However, in all previous investigations of epidemiological transient dynamics (Hosack et al., 2008; Mari et al., 2018, 2019, 2021; Trevisin et al., 2022) based on reactivity analysis, the Euclidean $\ell^2$-norm was typically applied instead. While the latter might be an inferior alternative when it comes to epidemiological applications (see Materials and Methods, main text), it also represents a very common choice in the context of reactivity analysis and its extensions (Neubert and Caswell, 1997; Caswell and Neubert, 2005; Mari et al., 2017). In the following, for the sake of reference, we report the essential technical details and the main results of epidemicity analysis for the examples considered in the main text, this time applying the $\ell^2$-norm. To distinguish the metrics obtained with this norm from those reported in the main text, we identify them with a "$\ell^2$" superscript.

#### Discrete epidemicity index

The maximum one-step growth of any perturbation to an asymptotically stable DFE is given by

$$\mathcal{E}^{\ell^2} = \max_{||\mathbf{I}(0)||_2 \neq 0} \frac{||\mathbf{I}(1)||_2}{||\mathbf{I}(0)||_2} = \max_{||\mathbf{I}(0)||_2 \neq 0} \frac{||\mathbf{L}\mathbf{I}(0)||_2}{||\mathbf{I}(0)||_2} = \nu_{\max}(\mathbf{L}) \; ,$$

where $\nu_{\max}(\mathbf{L})$ is the largest singular value of the projection matrix (Horn and Johnson, 2012). Analytical results can be obtained only under the simplifying hypotheses that the rate of exit from the infected compartment does not vary much with respect to the age of infection ($\sigma_0 = \exp(-\gamma/2)$ and $\sigma_j = \exp(-\gamma) = \sigma$ for $j \in \{1, \ldots, q-1\}$), as assumed also in the main text. In this case, in fact, the projection matrix takes the form of a Frobenius companion matrix, for which singular values can be evaluated analytically (Kittaneh, 1995; Linden, 1998). In particular, we have

$$\mathcal{E}^{\ell^2} = \sqrt{\sigma^2 + \mathcal{R}_c^2 \sigma_0^2 \sum_{j=1}^{q-1} f_j^2} \; ,$$

with transient epidemic outbreaks being possible if $\mathcal{E}^{\ell^2} > 1$. The maximum one-step growth is achieved by a perturbation whose components are proportional to the right singular vector associated with the largest singular value of the projection matrix (Caswell and Neubert, 2005). Note that, if a more refined description of the exit from the infected compartment is needed, short-term epidemicity and all the related metrics can still be evaluated numerically.

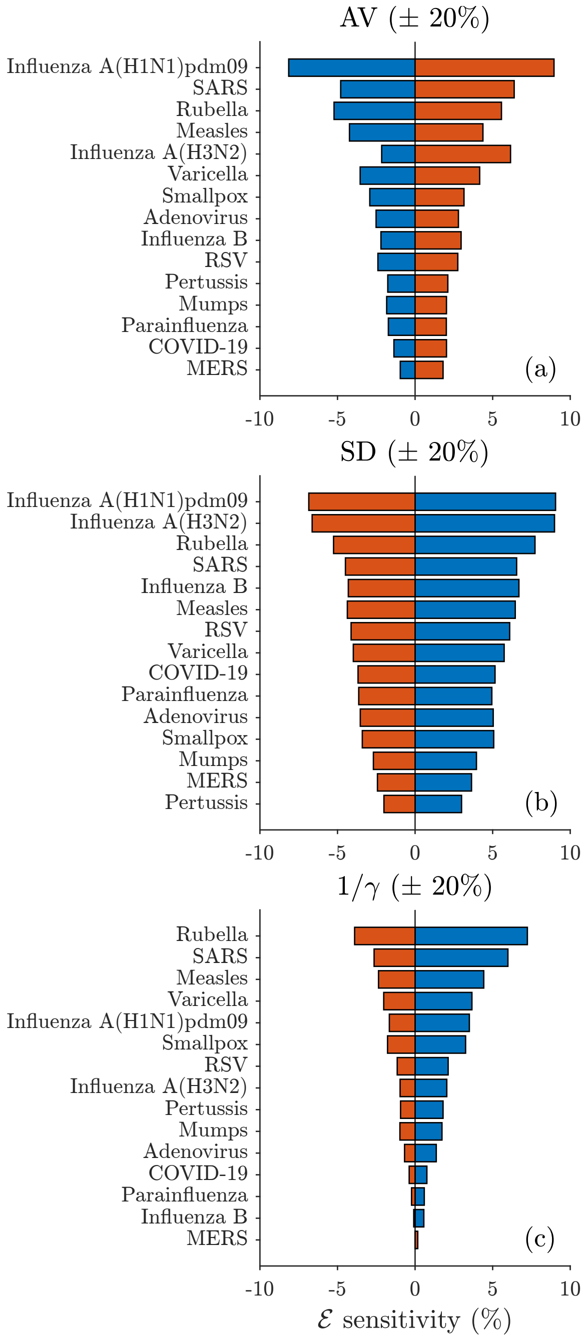


Figure S3: Sensitivity analysis of the $\mathcal{E}$ metric with respect to changes of the model parameterization. Sensitivity ranges are evaluated for the respiratory viruses listed in Table 1 (main text) by changing one at a time the relevant model parameters: (a) average generation time; (b) standard deviation of the generation time distribution; (c) infection duration. Note that $\mathcal{R}_0$ is not considered because its value does not affect the $\mathcal{E}$ metric. In all cases, $\pm 20\%$ parameter variations are considered, and sensitivity values are calculated with reference to the baseline values of $\mathcal{E}$ shown in Figure 3 (main text). Blue/red bars are associated with negative/positive variations of the considered parameter. Infections are ranked in decreasing order of the total lengths of their sensitivity bars (tornado diagram). Baseline parameter values as in Table 1 in the main text. See Section S2 for an extended description of the results shown in this figure.

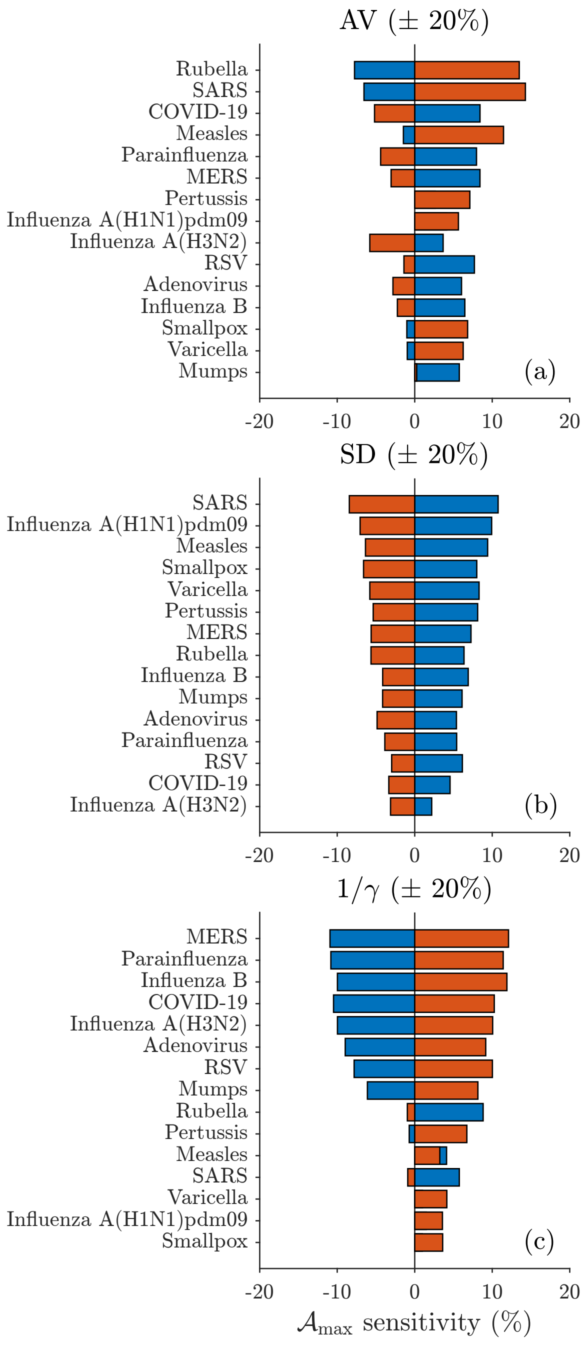


Figure S4: Sensitivity analysis of the $\mathcal{A}_{\max}$ metric with respect to changes of the model parameterization. Details as in Figure S3.

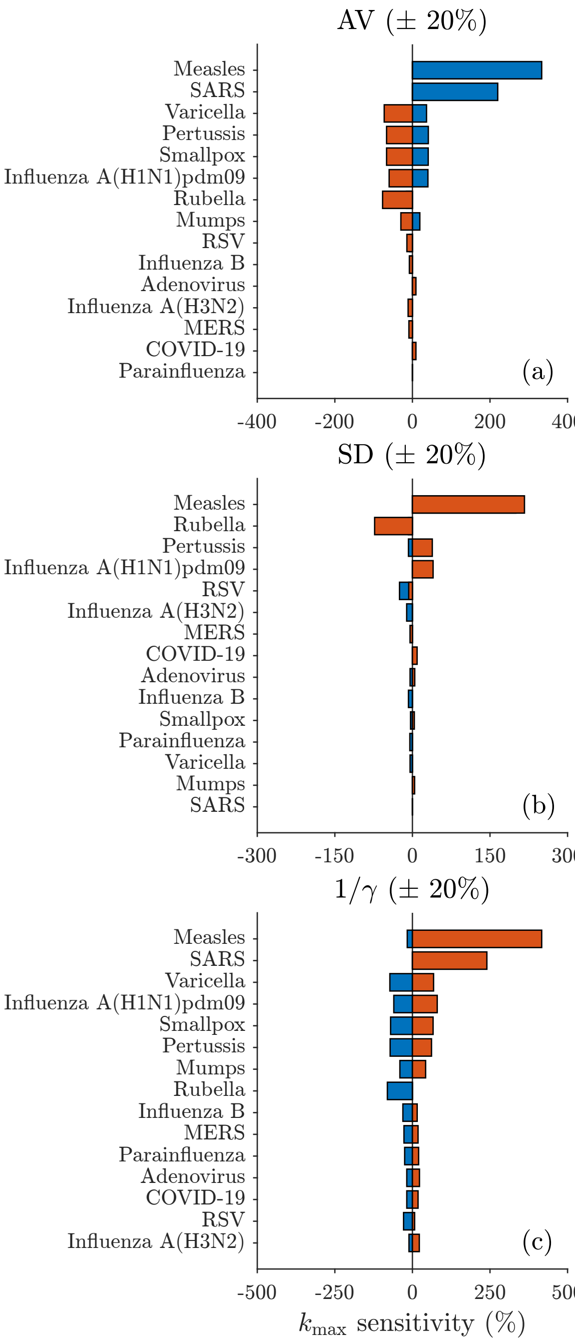


Figure S5: Sensitivity analysis of the $k_{\max}$ metric with respect to changes of the model parameterization. Details as in Figure S3.

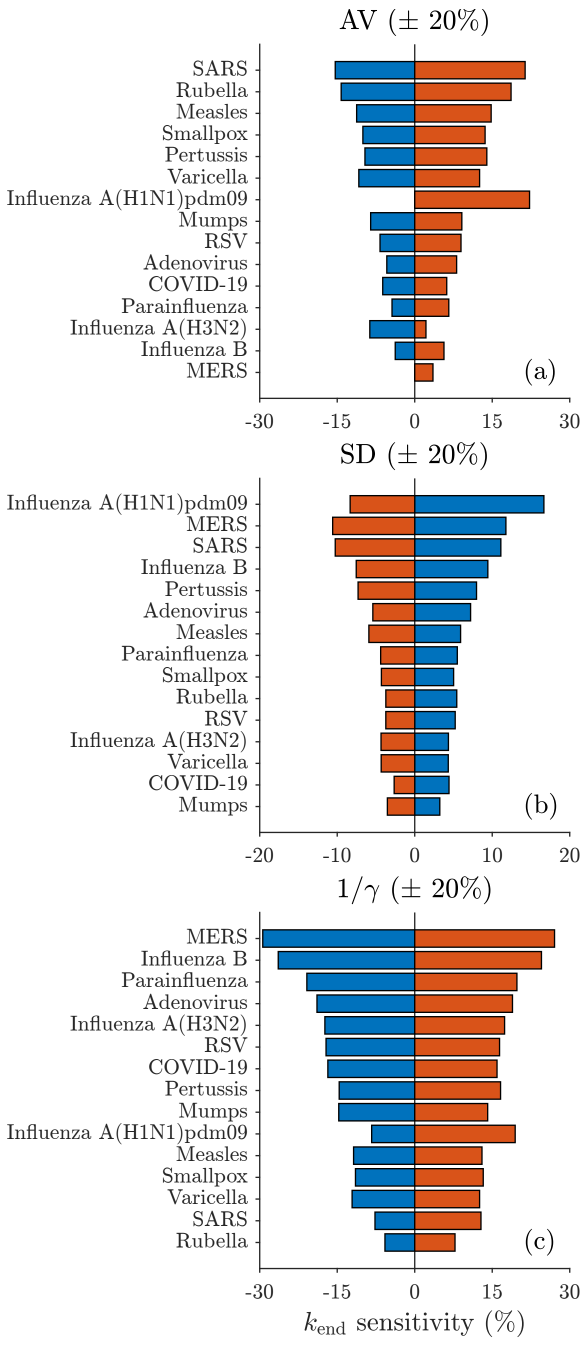


Figure S6: Sensitivity analysis of the $k_{\mathrm{end}}$ metric with respect to changes of the model parameterization. Details as in Figure S3.

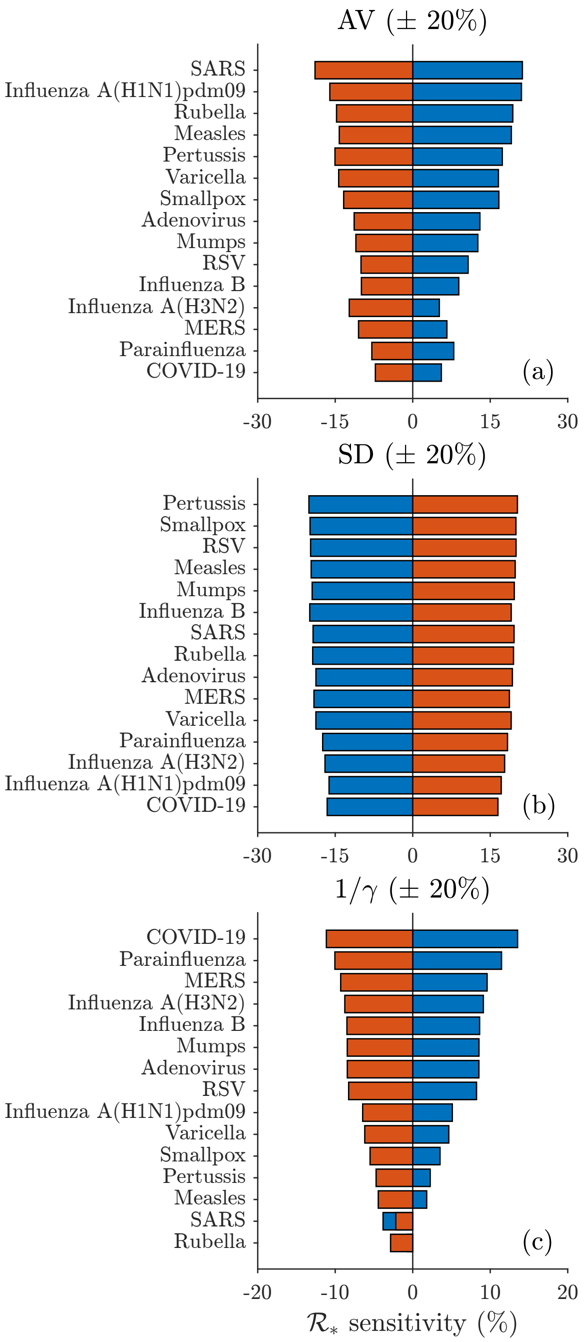


Figure S7: Sensitivity analysis of the $\mathcal{R}_*$ metric with respect to changes of the model parameterization. Details as in Figure S3.

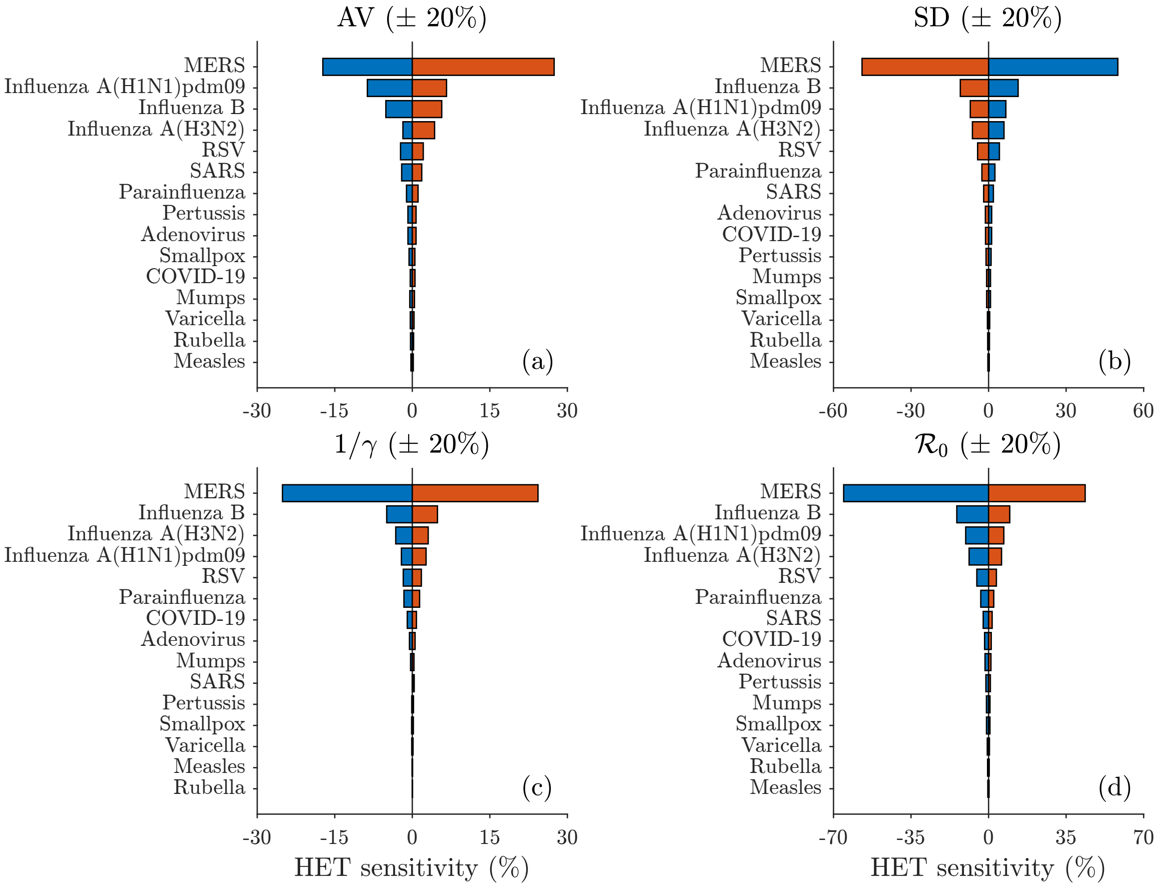


Figure S8: Sensitivity analysis of the HET metric with respect to changes of the model parameterization. Details as in Figure S3. Note that, for this metric, the influence of $\mathcal{R}_0$ has also been considered.

#### RN epidemicity threshold

Imposing $\mathcal{E}^{\ell 2} = 1$ in the previous equation, we obtain

$$\mathcal{R}_*^{\ell 2} = \frac{1}{\sigma_0}\sqrt{\frac{1-\sigma^2}{\sum_j f_j^2}}\,.$$

Therefore, the condition on the control RN to be imposed in order to prevent short-term epidemicity is RN $< \mathcal{R}_*^{\ell 2}$.

#### Herd epidemicity threshold

If information on a pathogen's $\mathcal{R}_0$ is also available, the minimum share of the population to be involved in preventive containment measures in order to avoid short-term epidemicity can be evaluated as

$$\mathrm{HET}^{\ell 2} = \max\left(0, 1 - \frac{\mathcal{R}_*^{\ell 2}}{\mathcal{R}_0}\right)\,.$$

#### Temporal evolution of transient outbreaks

The amplification envelope is defined as

$$\mathcal{A}^{\ell 2}(k) = \max_{||\mathbf{I}(0)||_2 \neq 0} \frac{||\mathbf{I}(k)||_2}{||\mathbf{I}(0)||_2} = \max_{||\mathbf{I}(0)||_2 \neq 0} \frac{||\mathbf{L}^k \mathbf{I}(0)||_2}{||\mathbf{I}(0)||_2}$$

and can be evaluated as the largest singular value of matrix $\mathbf{L}^k$ (Horn and Johnson, 2012). The largest amplification at time $k$ is achieved by a perturbation whose structure is proportional to the right singular value corresponding to the largest singular value of $\mathbf{L}^k$ (Caswell and Neubert, 2005). The epidemicity metrics based on the features of the amplification envelope (namely: the maximum amplification achieved by any perturbation at any time, $\mathcal{A}_{\max}^{\ell 2}$; the peak time of the amplification envelope, $k_{\max}^{\ell 2}$; and the time at which the maximum amplification becomes smaller than the initial perturbation in the chosen norm, $k_{\mathrm{end}}^{\ell 2}$) can be obtained by repeated applications of the projection matrix and have corresponding definitions to those provided with reference to the $\ell^1$-norm (Materials and Methods, main text).

#### Epidemicity analysis for viral respiratory infectious diseases

The epidemicity analysis of 15 infections caused by respiratory viruses reported in Figure 3 in the main text ($\ell^1$-norm) can be repeated here using the $\ell^2$-norm. Note that, while for any finite-sized state vector $\mathbf{w}$ the Cauchy–Schwarz inequality imposes $||\mathbf{w}||_1 \geq ||\mathbf{w}||_2$ (Horn and Johnson, 2012), it is not possible to infer a priori any relationship between the epidemicity (or, more generally, reactivity) properties evaluated with different norms (Harrington et al., 2022).

The results obtained in this case are qualitatively similar to those obtained with the $\ell^1$-norm, yet quantitatively quite different, as shown in Figure S9). In particular, for each disease, the one-step maximum amplification ($\mathcal{E}^{\ell^2}$) and the maximum amplification overall ($\mathcal{A}^{\ell^2}_{\max}$) evaluated with the $\ell^2$-norm are both smaller than their respective counterparts computed with the $\ell^1$-norm ($\mathcal{E}$ and $\mathcal{A}_{\max}$). As a consequence, using the $\ell^2$-norm, the estimate of the control RN that must be imposed to avoid short-term epidemicity is higher than it is with the $\ell^1$-norm ($\mathcal{R}^{\ell^2}_* > \mathcal{R}_*$); in contrast, the opposite is true when considering the herd epidemicity threshold ($\text{HET}^{\ell^2} < \text{HET}$). Moreover, for each disease, outbreaks are estimated to be shorter when the $\ell^2$-norm is used, in terms of both the time to the prevalence peak ($k^{\ell^2}_{\max} < k_{\max}$) and the maximum duration of the outbreak ($k^{\ell^2}_{\text{end}} < k_{\text{end}}$). While comparing the results obtained with the two different norms may not be particularly informative, the sheer differences existing between the two cases reinforce the notion that the choice of a suitable, epidemiologically motivated norm is an important preliminary step to performing an effective analysis of transient outbreaks and to reducing the risk of under/overestimating short-term epidemicity.

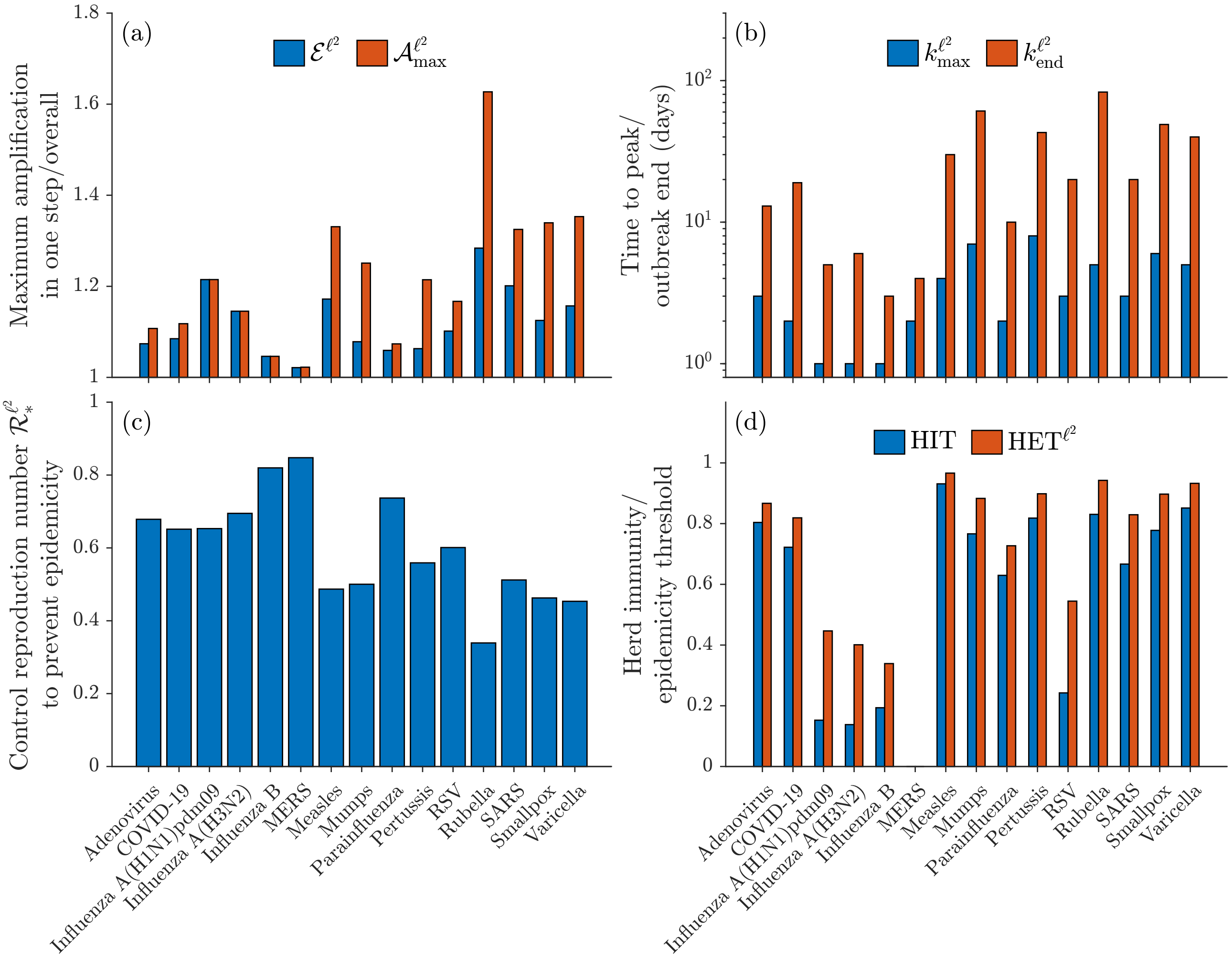


Figure S9: Summary of the transient epidemicity results for different respiratory viruses obtained with the $\ell^2$-norm. Note that, in panel (d), both the blue and the red bars for MERS are missing because $\mathcal{R}_0 < 1$ and $\mathcal{R}^{\ell^2}_* > \mathcal{R}_0$. Details as in Figure 3 in the main text, except for the use of a different norm to evaluate transient amplifications of the perturbations to the DFE. See Section S3 for an extended description of the results shown in this figure.

Gani, R. and Leach, S. (2001). Transmission potential of smallpox in contemporary populations. *Nature*, 414(6865):748–751.

Ganyani, T., Kremer, C., Chen, D., Torneri, A., Faes, C., Wallinga, J., and Hens, N. (2020). Estimating the generation interval for coronavirus disease (COVID-19) based on symptom onset data, March 2020. *Eurosurveillance*, 25(17):2000257.

Garnett, G. and Grenfell, B. (1992). The epidemiology of varicella–zoster virus infections: a mathematical model. *Epidemiology & Infection*, 108(3):495–511.

Gatto, M., Bertuzzo, E., Mari, L., Miccoli, S., Carraro, L., Casagrandi, R., and Rinaldo, A. (2020). Spread and dynamics of the COVID-19 epidemic in Italy: Effects of emergency containment measures. *Proceedings of the National Academy of Sciences*, 117(19):10484–10491.

Guo, Z., Tong, L., Xu, S., Li, B., Wang, Z., and Liu, Y. (2020). Epidemiological analysis of an outbreak of an adenovirus type 7 infection in a boot camp in China. *PloS One*, 15(6):e0232948.

Harrington, P. D., Lewis, M. A., and van den Driessche, P. (2022). Reactivity, attenuation, and transients in metapopulations. *SIAM Journal on Applied Dynamical Systems*, 21(2):1287–1321.

Hethcote, H. W. (1997). An age-structured model for pertussis transmission. *Mathematical Biosciences*, 145(2):89–136.

Hogan, A. B., Glass, K., Moore, H. C., and Anderssen, R. S. (2016). Exploring the dynamics of respiratory syncytial virus (RSV) transmission in children. *Theoretical Population Biology*, 110:78–85.

Horn, R. A. and Johnson, C. R. (2012). *Matrix Analysis*. Cambridge University Press.

Hosack, G. R., Rossignol, P. A., and van den Driessche, P. (2008). The control of vector-borne disease epidemics. *Journal of Theoretical Biology*, 255(1):16–25.

Kittaneh, F. (1995). Singular values of companion matrices and bounds on zeros of polynomials. *SIAM Journal on Matrix Analysis and Applications*, 16(1):333–340.

Kucharski, A. and Althaus, C. L. (2015). The role of superspreading in Middle East respiratory syndrome coronavirus (MERS-CoV) transmission. *Eurosurveillance*, 20(25):21167.

Lehtinen, S., Ashcroft, P., and Bonhoeffer, S. (2021). On the relationship between serial interval, infectiousness profile and generation time. *Journal of the Royal Society Interface*, 18(174):20200756.

Levy, J. W., Cowling, B. J., Simmerman, J. M., Olsen, S. J., Fang, V. J., Suntarattiwong, P., Jarman, R. G., Klick, B., and Chotipitayasunondh, T. (2013). The serial intervals of seasonal and pandemic influenza viruses in households in Bangkok, Thailand. *American Journal of Epidemiology*, 177(12):1443–1451.

Li, Y., Liu, X., and Wang, L. (2018). Modelling the transmission dynamics and control of mumps in mainland China. *International Journal of Environmental Research and Public Health*, 15(1):33.

Linden, H. (1998). Bounds for the zeros of polynomials from eigenvalues and singular values of some companion matrices. *Linear Algebra and its Applications*, 271(1-3):41–82.

Lipsitch, M., Cohen, T., Cooper, B., Robins, J. M., Ma, S., James, L., Gopalakrishna, G., Chew, S. K., Tan, C. C., Samore, M. H., et al. (2003). Transmission dynamics and control of severe acute respiratory syndrome. *Science*, 300(5627):1966–1970.

Mari, L., Casagrandi, R., Bertuzzo, E., Pasetto, D., Miccoli, S., Rinaldo, A., and Gatto, M. (2021). The epidemicity index of recurrent SARS-CoV-2 infections. *Nature Communications*, 12(1):2752.

Mari, L., Casagrandi, R., Bertuzzo, E., Rinaldo, A., and Gatto, M. (2019). Conditions for transient epidemics of waterborne disease in spatially explicit systems. *Royal Society Open Science*, 6(5):181517.

Mari, L., Casagrandi, R., Rinaldo, A., and Gatto, M. (2017). A generalized definition of reactivity for ecological systems and the problem of transient species dynamics. *Methods in Ecology and Evolution*, 8(11):1574–1584.

Mari, L., Casagrandi, R., Rinaldo, A., and Gatto, M. (2018). Epidemicity thresholds for water-borne and water-related diseases. *Journal of Theoretical Biology*, 447:126–138.

Neubert, M. G. and Caswell, H. (1997). Alternatives to resilience for measuring the responses of ecological systems to perturbations. *Ecology*, 78(3):653–665.

Reis, J. and Shaman, J. (2018). Simulation of four respiratory viruses and inference of epidemiological parameters. *Infectious Disease Modelling*, 3:23–34.

Rui, J., Wang, Q., Lv, J., Zhao, B., Hu, Q., Du, H., Gong, W., Zhao, Z., Xu, J., Zhu, Y., et al. (2022). The transmission dynamics of Middle East respiratory syndrome coronavirus. *Travel Medicine and Infectious Disease*, 45:102243.

Sato, M., Hosoya, M., Kato, K., and Suzuki, H. (2005). Viral shedding in children with influenza virus infections treated with neuraminidase inhibitors. *The Pediatric infectious disease journal*, 24(10):931–932.

Tang, X., Zhao, S., Chiu, A. P., Ma, H., Xie, X., Mei, S., Kong, D., Qin, Y., Chen, Z., Wang, X., et al. (2017). Modelling the transmission and control strategies of varicella among school children in Shenzhen, China. *PLoS One*, 12(5):e0177514.

Thompson, K. M. (2016). Evolution and use of dynamic transmission models for measles and rubella risk and policy analysis. *Risk Analysis*, 36(7):1383–1403.

Trentini, F., Pariani, E., Bella, A., Diurno, G., Crottogini, L., Rizzo, C., Merler, S., and Ajelli, M. (2022). Characterizing the transmission patterns of seasonal influenza in Italy: lessons from the last decade. *BMC Public Health*, 22(1):1–9.

Trevisin, C., Lemaitre, J. C., Mari, L., Pasetto, D., Gatto, M., and Rinaldo, A. (2022). Epidemicity of cholera spread and the fate of infection control measures. *Journal of the Royal Society Interface*, 19(188):20210844.

van den Driessche, P. (2017). Reproduction numbers of infectious disease models. *Infectious Disease Modelling*, 2(3):288–303.

Vink, M. A., Bootsma, M. C. J., and Wallinga, J. (2014). Serial intervals of respiratory infectious diseases: a systematic review and analysis. *American Journal of Epidemiology*, 180(9):865–875.

Wang, B., Peng, M., Yang, L., Li, G., Yang, J., Yundan, C., Zeng, X., Wei, Q., Han, Q., Liu, C., et al. (2021). Clinical and immunological characteristics of patients with adenovirus infection at different altitude areas in tibet, china. *Frontiers in Cellular and Infection Microbiology*, 11:739429.

Wang, L., Chu, C., Yang, G., Hao, R., Li, Z., Cao, Z., Qiu, S., Li, P., Wu, Z., Yuan, Z., et al. (2014). Transmission characteristics of different students during a school outbreak of (H1N1) pdm09 influenza in China, 2009. *Scientific Reports*, 4(1):1–8.

Zhang, W., Ma, X., Zhang, Y., and Luo, X. (2022). Dynamical models of acute respiratory illness caused by human adenovirus on campus. *Frontiers in Physics*, 10:1325.
